# Phase 1 trial of a Candidate Recombinant Virus-Like Particle Vaccine for Covid-19 Disease Produced in Plants

**DOI:** 10.1101/2020.11.04.20226282

**Authors:** Brian J Ward, Philipe Gobeil, Annie Séguin, Judith Atkins, Iohann Boulay, Pierre-Yves Charbonneau, Manon Couture, Marc-André D’Aoust, Jiwanjeet Dhaliwall, Carolyn Finkle, Karen Hager, Asif Mahmood, Alexander Makarkov, Matthew Cheng, Stéphane Pillet, Patricia Schimke, Sylvie St-Martin, Sonia Trépanier, Nathalie Landry

**Author notes:** Corresponding author*: Nathalie Landry, 1020 Route de l’Église, Bureau 600, Québec, Qc, Canada, G1V 3V9.

## Abstract

**Background:** The stabilized prefusion form of the SARS-CoV-2 spike protein is produced by transient expression in *Nicotiana benthamiana*. The trimeric spike glycoproteins are displayed at the surface of self-assembling Virus-Like-Particles that mimic the shape and the size of the virus. The candidate vaccine (CoVLP) administered alone or with AS03 or CpG1018 adjuvants was evaluated in a Phase 1 trial in healthy adults. (ClinicalTrials.gov number NCT04450004)

**Methods:** The study was a randomized, partially-blinded, prime-boost 21 days apart, dose-escalation Phase 1 study intended to assess the safety, tolerability, and immunogenicity of CoVLP at three dose levels (3.75 µg, 7.5 µg, and 15 µg) unadjuvanted or adjuvanted with either CpG 1018 or AS03 in 180 SARS-CoV-2 seronegative healthy adults 18 to 55 years of age. Enrollment was staggered for dose-escalation. At each dose level, the vaccine was initially administered to a small number of subjects. Vaccination of the remaining subjects at the same dose level and the next higher vaccine dose level was administered with approval of an Independent Data Monitoring Committee (IDMC). The same procedure was followed for the second vaccine administration. Monitoring of safety signals was performed throughout the study with pre-determined pausing/stopping rules if there was clear evidence of harmful effects such as severe adverse events (AEs) related to the treatment. The primary endpoints were the safety and tolerability of the vaccine after each dose and the immunogenicity as assessed by neutralizing antibody responses assessed using a vesicular stomatitis virus (VSV) pseudovirion assay and interferon-gamma (IFN-γ) and interleukin-4 (IL-4) ELISpot assays at Days 0, 21 and 42. Secondary endpoints were anti-spike antibody responses by ELISA and neutralizing antibodies measured by live virus plaque reduction neutralization test (PRNT) assay at Days 0, 21 and 42 and immunogenicity with additional safety and immunogenicity endpoints planned for 6-months following the last vaccination. The anti-spike and neutralizing antibody responses were compared with 23 convalescent serum samples from symptomatic Covid-19 patients. We performed a primary analysis at day 42.

**Results:** A total of 180 subjects (102 females: 78 males: average 34.3 years) were recruited to the study and interim safety and immunogenicity data up to day 42 after the first dose are reported here. There was no obvious CoVLP dose effect in safety outcomes for any of the formulations tested and all formulations were generally well-tolerated. Most solicited local and systemic AEs were mild-moderate and transient. Reactogenicity was increased in all adjuvanted formulations and was generally highest in the CoVLP+AS03 groups. Local and systemic adverse events were reported with similar frequency after the first and second doses in subjects who received either CoVLP alone or CoVLP+CpG1018 but increased in both frequency and severity after the second dose in the CoVLP+AS03 groups. CoVLP alone only elicited a weak total anti-spike IgG response at the highest dose level and little-to-no neutralization antibody response, even after the second dose. Cellular responses in the CoVLP alone groups (IFNγ and IL-4) were detectable after the second dose but were still only marginally above background levels. The addition of either adjuvant substantially increased both antibody and cellular responses at most CoVLP dose levels and changes were most pronounced after the second dose. However, a substantial neutralizing antibody response after the first dose was only seen in all CoVLP+AS03 groups. After the second dose, both total anti-spike IgG and neutralizing antibody titers in the CoVLP+AS03 groups were higher than those in the CoVLP+CpG1018 groups. The antibody titers achieved were either similar to (CoVLP+CpG1018) or at least 10-times higher (CoVLP+AS03) than those seen in convalescent plasma. Administration of CoVLP with either adjuvant also significantly increased the cellular responses. After 2 doses, both IFN-γ and IL-4 responses were significantly increased in the CoVLP+CpG1018 groups. In the CoVLP+AS03 groups, significant increases in the cellular responses were observed after the first dose while IFN-γ and IL-4 increased further in both magnitude and number of subjects responding after the second dose. Again, the cellular responses in the CoVLP+AS03 groups were higher than those seen in the CoVLP+CpG1018 groups.

**Conclusion:** These data demonstrate that CoVLP administered with either CpG1018 or AS03 has a safety profile similar to other candidate vaccines for SARS-CoV-2. When administered with either AS03 or CpG1018, several of the CoVLP dose levels elicited strong humoral and T cell responses after the second dose. When administered with AS03, even the 3.75 μg CoVLP dose elicited neutralizing antibody titers that were ∼10-times higher than those observed in individuals recovering from Covid-19 as well as consistent and balanced IFN-γ and IL-4 responses. Although many CoVLP formulations were immunogenic, in the absence of established correlates of protection and given the advantages of dose-sparing in the context of the on-going pandemic, these findings suggest that CoVLP (3.75 μg)+AS03 has a good benefit/risk ratio and support the transition of this formulation to studies in expanded populations and to efficacy evaluations

**Shorter Abstract:** *Background:* Virus-like particles (VLP) displaying recombinant SARS-CoV-2 spike protein trimers were produced by transient expression in *Nicotiana benthamiana*. This candidate vaccine (CoVLP) was evaluated in healthy adults 18-55 years of age alone or with AS03 or CpG1018 (NCT04450004).

*Methods:* This randomized, partially-blinded, two-dose, dose-escalation study assessed CoVLP (3.75, 7.5 or 15 µg/dose) administered intramuscularly alone or with CpG1018 or AS03 in SARS-CoV-2 seronegative adults (18-55 years). Primary endpoints of safety and immunogenicity are reported to day 42. Neutralizing antibodies (NtAb) were assessed using a VSV pseudovirus assay and cellular responses by ELISpot (IFNγ, IL-4).

*Results:* 180 subjects (avg.34.3yrs) were recruited. All formulations were well-tolerated but adjuvants increased reactogenicity. Adverse events were highest in the CoVLP+AS03 groups and increased in frequency/severity after dose two. CoVLP alone elicited weak humoral responses but modest cellular responses were detectable after dose two. Both adjuvants increased immunogenicity significantly, particularly after dose two. A significant NtAb response after dose one was only seen in CoVLP+AS03 groups. The vaccine dose had little impact on levels of NtAb responses achieved in the CoVLP+AS03 groups. Both adjuvants also increased IFNγ and IL-4 responses but these cellular responses also tended to be highest in the AS03-adjuvanted groups.

*Conclusion:* CoVLP ± adjuvants was well-tolerated. Several adjuvanted formulations elicited strong humoral and T cell responses after dose 2. Even at the lowest CoVLP+AS03 dose, NtAb titers were ∼10-times higher than in convalescent serum with a balanced IFNγ and IL-4 response. These findings support the transition of CoVLP (3.75μg+AS03) to further clinical evaluation.

**Research In Context:** *Evidence before this study:* The severe acute respiratory syndrome coronavirus 2 (SARS-CoV-2) was recognized as the causative agent of COVID-19 in early 2020. Since that time, >150 candidate vaccines are reported to be under development of which 47 have entered clinical trials (https://www.who.int/publications/m/item/draft-landscape-of-covid-19-candidate-vaccines accessed Nov 4, 2020). No vaccine to prevent COVID-19 has been licensed yet for either emergency or general use in North America or Europe. We searched PubMed for research articles published between July 2019 and November 4, 2020, using the terms “SARS-CoV-2”, “vaccine”, “clinical trial” OR “human”, AND “phase”. The same terms were used to search ClinTrials.gov. No language restrictions were applied. We identified 10 peer-reviewed studies, describing phase 1 or 1/2 trials using a range of novel (eg: RNA, DNA, non-replicating virus vectored) and more traditional vaccine approaches (eg: inactivated virus or recombinant protein ± adjuvants). None of these candidate vaccines was produced in plants. These reports demonstrate that several different vaccination strategies (typically delivered in two doses 14-28 days apart) are capable of eliciting neutralizing antibody responses. In several cases, vaccine-induced cellular responses against SARS-COV-2 antigens - predominantly the spike (S) protein - can also be demonstrated. Although local and systemic adverse events following vaccination have varied between reports, the trials published to date suggest that each of these candidate vaccines is well-tolerated in the context of an evolving pandemic.

*Added value of this study:* We report the results of the first clinical study of CoVLP, a virus-like particle (VLP) vaccine that is produced by transient transfection of *Nicotiana benthamiana* plants. These VLPs spontaneously assemble at the plant cell membrane and display SARS-COV-2 trimers of stabilized pre-fusion S protein on their surface. The vaccine was administered as two intramuscular doses 21 days apart at three dose levels (S protein content 3.75, 7,5 or 15μg) alone or adjuvanted with either CpG1018 or AS03. All formulations were well-tolerated although both adjuvants increased reactogenicity. Humoral (anti-S IgG and neutralizing antibodies) as well as cellular responses (IFNg and IL4 ELISpots) were detectable in almost all subjects who received adjuvanted formulations 21 days after the second dose at all COVLP dose levels. Both antibody and cellular responses were highest in subjects who received AS03-adjuvanted formulations. Even at the lowest dose level (3.75μg), the neutralizing antibody titers 21 days after the second dose in subjects who received the AS03-afdjuvanted vaccine were 10-50-fold higher than those seen in subjects recovering from COVID-19 infection.

*Implications of all the available evidence:* Effective vaccines against SARS-CoV-2 are urgently needed to reduce the burden of disease and contribute to ending the global pandemic. Although no immune correlates for SARS-CoV-2 have been defined, it is likely that both arms of the immune system contribute to protection. After two doses of CoVLP (3.75μg+AS03), strong humoral and cellular responses were induced supporting the further clinical development of this vaccine.

## Introduction

A novel coronavirus, Severe Acute Respiratory Syndrome Coronavirus 2 (SARS-CoV-2) jumped across a species barrier in China in late 2019 ^1,2^ and spread rapidly around the globe leading to the World Health Organization’s declaration of a pandemic on March 11, 2020 ^3^. As of October 11th, 2020, more than 37 million cases of Covid-19 have been reported with >1 million deaths ^4^. Although many therapeutic strategies has been tried ^5,6 7^, the current options remain limited in both number and efficacy. Simultaneously, there has been a massive global effort to develop vaccines. This effort was primed to some extent by prior experience with other highly pathogenic human coronaviruses, SARS and MERS^8^ and the far-sighted efforts of the Coalition for Epidemic Preparedness Innovations (CEPI) to develop vaccines for a ‘short-list’ of pathogens with pandemic potential ^9^. At the time of writing, >150 vaccine candidates have been announced, >30 are in clinical trials and a small number have been authorized for limited use in some countries ^10^. These candidates are based on a wide range of traditional and novel platforms including mRNA, DNA, inactivated virus, live viral vectors, recombinant proteins, peptides, and virus-like particles (VLPs) ^11,12^. No vaccine has been approved for general use yet in North America.

Adding to the complexity of this situation is the facts that no correlate of immunity has been defined for any highly pathogenic coronavirus ^13,14^ and that such correlates might differ between vaccines^15^. Nonetheless, a protective role for both humoral and cell-mediated immunity against coronaviruses has been suggested ^16,17^. Antibody responses against the spike (S) protein have the potential to protect from infection ^18-20^ and convalescent plasma with high titers of anti-S antibody have therapeutic benefit in selected patients ^7,21^. However, a substantial proportion of people who develop Covid-19 fail to generate antibodies, and data from the SARS-CoV-1 outbreak of 2002–2003 ^22^ and the current pandemic suggest that antibody responses can be short-lived, disappearing within months of infection in some patients^23^. In contrast, T cell immunity may be critical for recovery from Covid-19^24,25^ and was shown to persist for up to 11 years after SARS-CoV-1 infection ^26^. T cells can provide substantial protection in animal models of highly pathogenic coronavirus infection.

We report here the results of a Phase 1 study initiated in July 2020 evaluating the safety, tolerability and immunogenicity of two doses, 21-days apart of 3.75, 7.5 or 15 µg of a virus-like-particle vaccine candidate for Covid-19 produced in plants (hereafter called CoVLP). This recombinant platform has been used to produce hemagglutinin (HA)-bearing VLP vaccines for avian (monovalent) and seasonal (quadrivalent) influenza that induce balanced humoral and T cell responses ^28-31^. The CoVLP vaccine was administered alone or with AS03 or CpG1018 adjuvants in healthy adults 18-55 years of age.

## Methods

### The CoVLP Vaccine and Adjuvants

The full-length S glycoprotein of SARS-CoV-2, strain hCoV-19/USA/CA2/2020, corresponding in sequence to nucleotides 21563 to 25384 from EPI_ISL_406036 in GISAID database (https://www.gisaid.org/) was expressed in *Nicotiana benthamiana* plants as previously described ^32^. The S protein was modified with R667G, R668S and R670S substitutions at the S1/S2 cleavage site to increase stability, and K971P and V972P substitutions to stabilize the protein in prefusion conformation. The signal peptide was replaced with a plant gene signal peptide and the transmembrane domain (TM) and cytoplasmic tail (CT) of S protein was also replaced with TM/CT from Influenza H5 A/Indonesia/5/2005 to increase VLP assembly and budding. The self-assembled VLPs bearing S protein trimers were isolated from the plant matrix and subsequently purified using a process similar to that described for the influenza vaccine candidates ^28^.

The AS03 adjuvant, an oil-in-water emulsion containing tocopherol and squalene, was supplied by GlaxoSmithKline. The CpG 1018 adjuvant, composed of cytosine phosphoguanine (CpG) motifs, was supplied by Dynavax. The CoVLP vaccine and adjuvants were mixed immediately prior to administration

### Study design

The Phase 1 study was conducted at two sites in Quebec City and Montreal (Protocol available in Supplemental Material). At screening, health status was assessed by medical history, physical exam and clinical laboratory findings including detection of antibodies to SARS-CoV-2 (ElecSys: Roche Diagnostics). Healthy seronegative subjects 18-55 years of age who met all inclusion criteria and no exclusion criterion were enrolled and randomized into nine groups. Randomised subjects received two doses, 21 days apart of CoVLP at doses of 3.75, 7.5 or 15 µg unadjuvanted or adjuvanted with AS03 or CpG1018. The participants and the personnel collecting the safety information and testing laboratories remained blinded to treatment allocation. On Day 0 (D0: pre-first dose), D21 (pre-second dose) and D42 (post second dose), serum and peripheral blood mononuclear cells (PBMC) were processed for immune outcomes as described previously ^28^.

### Safety

For details of safety monitoring, see the Protocol in Suppl. Materials. Briefly, enrollment was staggered for dose-escalation with sentinel subjects at each dose level (n=6) and independent data monitoring committee (IDMC) review of D3 safety data at 10% and 30% recruitment before each dose acceleration. The same process was followed for the second vaccine administration. Monitoring of safety signals was performed throughout the study (Suppl Material: pp 2). Solicited adverse events (AEs) were assessed by the subjects as Grade 1 to 4 (mild, moderate, severe, or potentially life-threatening). Unsolicited AEs, and AEs leading to subject withdrawal were collected up to D21 after each vaccination. All serious AEs (SAE), Adverse Events of Special Interest (AESIs), and pregnancies will be collected for 6 months. Potential cases of vaccine enhanced disease (VED), hypersensitivity and potential immune-mediated diseases (pIMDs) are being monitored throughout the study (see Suppl. Material: pp 7, Tables S2, S3).

### Immunogenicity assessments

The primary immunological outcomes were neutralizing antibody (NAb) responses measured using a VSV pseudovirion assay (Nexelis Inc, Laval, QC) and IFNγ and IL-4 cellular responses measured by ELISpot at D0, 21 and 42. Secondary immunological outcomes were total anti-spike IgG responses by ELISA and NAb responses by plaque reduction neutralization test (PRNT: Vismedri S.r.l., Siena, Italy) Details of these assays are provided in the Suppl. Material (pp 3-5).

Antibody responses were compared with those of individuals who had recovered from PCR-confirmed Covid-19 obtained from Solomon Park (Burien, WA)), Sanguine BioSciences (Sherman Oaks, CA) and M Cheng (McGill University Health Center, Montreal, Quebec). The severity of disease ranged from mild-moderate (n=23) to severe/critical (n=11) (Suppl. Material: Table S1 for patient characteristics: pp 6).

### Analysis Populations and Statistical Analysis Plan

Overall, 180 healthy SATRS-COV-2 seronegative male and female subjects 18 to 55 years of age were randomized in a 1:1:1:1:1:1:1:1:1 ratio into nine treatment groups. The sample size made it possible to perform the initial evaluation of the vaccine immunogenicity and detect major differences in rates of AEs between groups. The sample size was not large enough to detect all types, including less frequent or rare, AEs.

The analyses of all immunogenicity endpoints were based on the Per Protocol set (PP) and are described in the Statistical Analysis Plan provided as Supplementary Material.

## Results

### Demographic and baseline clinical characteristics

Participant demographics are presented in Table 1 and subject disposition up to D42 is presented in Table 2 and in Suppl. Materials: Fig. S1: pp. 19. More women (56.7%) than men (43.3%) were enrolled but the female:male ratio was the same for each CoVLP dose level and in each of the overall groups (unadjuvanted CoVLP, CoVLP+AS03, CoVLP+CpG1018). Subjects were mostly White (96%) with 2% each of Black or African American and Asian subjects. The average age was 34.3 years. A total of 180 subjects received the first dose of vaccine and 178 subjects received both doses. See Fig. S1 in Suppl. Materials for details.

**Table 1:** Summary of demographics and baseline characteristics (NCT04450004)

|  | CoVLP 3.75 µg |  |  | CoVLP 7.5 µg |  |  | CoVLP 15 µg |  |  | All |
| --- | --- | --- | --- | --- | --- | --- | --- | --- | --- | --- |
|  | Unadjuvanted | Adjuvanted<br>with CpG<br>1018 | Adjuvanted<br>with AS03 | Unadjuvanted | Adjuvanted<br>with CpG<br>1018 | Adjuvanted<br>with AS03 | Unadjuvanted | Adjuvanted<br>with CpG<br>1018 | Adjuvanted<br>with AS03 |  |
| Subjects | 20 | 20 | 20 | 20 | 20 | 20 | 20 | 20 | 20 | 180 |
| Sex, n (%) |  |  |  |  |  |  |  |  |  |  |
| Male | 9 (45.0) | 10 (50.0) | 5 (25.0) | 10 (50.0) | 8 (40.0) | 8 (40.0) | 7 (35.0) | 10 (50.0) | 11 (55.0) | 78 (43.3) |
| Female | 11 (55.0) | 10 (50.0) | 15 (75.0) | 10 (50.0) | 12 (60.0) | 12 (60.0) | 13 (65.0) | 10 (50.0) | 9 (45.0) | 102 (56.7) |
| Race, n (%) |  |  |  |  |  |  |  |  |  |  |
| White | 18 (90.0) | 20 (100.0) | 20 (100.0) | 20 (100.0) | 18 (90.0) | 19 (95.0) | 20 (100.0) | 18 (90.0) | 19 (95.0) | 172 (95.6) |
| Black or African<br>American | 1 (5.0) | 0 (0.0) | 0 (0.0) | 0 (0.0) | 1 (5.0) | 0 (0.0) | 0 (0.0) | 1 (5.0) | 1 (5.0) | 4 (2.2) |
| Asian | 1 (5.0) | 0 (0.0) | 0 (0.0) | 0 (0.0) | 1 (5.0) | 1 (5.0) | 0 (0.0) | 1 (5.0) | 0 (0.0) | 4 (2.2) |
| Ethnicity, n (%) |  |  |  |  |  |  |  |  |  |  |
| Hispanic/Latinx | 0 (0.0) | 2 (10.0) | 0 (0.0) | 1 (5.0) | 3 (15.0) | 1 (5.0) | 0 (0.0) | 1 (5.0) | 0 (0.0) | 9 (5.0) |
| Age at Vaccination |  |  |  |  |  |  |  |  |  |  |
| Mean ± SD | 34.9 ± 8.3 | 35.3 ± 11.0 | 34.7 ± 9.1 | 35.6 ± 8.0 | 32.4 ± 9.5 | 37.2 ± 7.8 | 34.1 ± 9.6 | 32.0 ± 9.0 | 32.7 ± 9.1 | 34.3 ± 9.0 |
| Median (range) | 35<br>(18-49) | 36<br>(18-53) | 36<br>(19-49) | 36<br>(20-50) | 31<br>(19-52) | 37<br>(21-55) | 31.5<br>(22-54) | 30<br>(19-51) | 32.5<br>(18-52) | 34<br>(18-55) |

SD: Standard Deviation

**Table 2:** Subject Disposition.

|  | CoVLP 3.75 µg |  |  | CoVLP 7.5 µg |  |  | CoVLP 15 µg |  |  | All |
| --- | --- | --- | --- | --- | --- | --- | --- | --- | --- | --- |
|  | Unadjuvanted<br>n (%) | Adjuvanted<br>with CpG<br>1018<br>n (%) | Adjuvanted<br>with AS03<br>n (%) | Unadjuvanted<br>n (%) | Adjuvanted<br>with CpG<br>1018<br>n (%) | Adjuvanted<br>with AS03<br>n (%) | Unadjuvanted<br>n (%) | Adjuvanted<br>with CpG<br>1018<br>n (%) | Adjuvanted<br>with AS03<br>n (%) |  |
| Screened Subjects |  |  |  |  |  |  |  |  |  | 413 |
| Screened Failure |  |  |  |  |  |  |  |  |  | 233 |
| Randomized Subjects | 20 | 20 | 20 | 20 | 20 | 20 | 20 | 20 | 20 | 180 |
| Vaccinated at Day 0 | 20 (100.0) | 20 (100.0) | 20 (100.0) | 20 (100.0) | 20 (100.0) | 20 (100.0) | 20 (100.0) | 20 (100.0) | 20 (100.0) | 180 (100.0) |
| Vaccinated at Day 21 | 20 (100.0) | 20 (100.0) | 19 (95.0) | 20 (100.0) | 20 (100.0) | 20 (100.0) | 20 (100.0) | 19 (95.0) | 20 (100.0) | 178 (98.9) |
| Safety Analysis Set | 20 (100.0) | 20 (100.0) | 20 (100.0) | 20 (100.0) | 20 (100.0) | 20 (100.0) | 20 (100.0) | 20 (100.0) | 20 (100.0) | 180 (100.0) |
| Per Protocol Set |  |  |  |  |  |  |  |  |  |  |
| Day 21 | 20 (100.0) | 20 (100.0) | 20 (100.0) | 20 (100.0) | 20 (100.0) | 20 (100.0) | 20 (100.0) | 19 (95.0) | 20 (100.0) | 179 (99.4) |
| Day 42 | 20 (100.0) | 20 (100.0) | 20 (100.0) | 20 (100.0) | 19 (95.0) | 20 (100.0) | 20 (100.0) | 19 (95.0) | 20 (100.0) | 178 (98.9) |
| Completed Through Day 21 | 20 (100.0) | 20 (100.0) | 20 (100.0) | 20 (100.0) | 20 (100.0) | 20 (100.0) | 20 (100.0) | 19 (95.0) | 20 (100.0) | 179 (99.4) |
| Early Withdrawal from Study | 0 (0.0) | 0 (0.0) | 0 (0.0) | 0 (0.0) | 0 (0.0) | 0 (0.0) | 0 (0.0) | 1 (5.0) | 0 (0.0) | 1 (0.6) |
| Adverse Event | 0 (0.0) | 0 (0.0) | 0 (0.0) | 0 (0.0) | 0 (0.0) | 0 (0.0) | 0 (0.0) | 0 (0.0) | 0 (0.0) | 0 (0.0) |
| Death | 0 (0.0) | 0 (0.0) | 0 (0.0) | 0 (0.0) | 0 (0.0) | 0 (0.0) | 0 (0.0) | 0 (0.0) | 0 (0.0) | 0 (0.0) |
| Lost to | 0 (0.0) | 0 (0.0) | 0 (0.0) | 0 (0.0) | 0 (0.0) | 0 (0.0) | 0 (0.0) | 0 (0.0) | 0 (0.0) | 0 (0.0) |
| Follow-up |  |  |  |  |  |  |  |  |  |  |
| Physician | 0 (0.0) | 0 (0.0) | 0 (0.0) | 0 (0.0) | 0 (0.0) | 0 (0.0) | 0 (0.0) | 0 (0.0) | 0 (0.0) | 0 (0.0) |
| Decision |  |  |  |  |  |  |  |  |  |  |
| Pregnancy | 0 (0.0) | 0 (0.0) | 0 (0.0) | 0 (0.0) | 0 (0.0) | 0 (0.0) | 0 (0.0) | 0 (0.0) | 0 (0.0) | 0 (0.0) |
| Protocol | 0 (0.0) | 0 (0.0) | 0 (0.0) | 0 (0.0) | 0 (0.0) | 0 (0.0) | 0 (0.0) | 0 (0.0) | 0 (0.0) | 0 (0.0) |
| Deviation |  |  |  |  |  |  |  |  |  |  |
| Site | 0 (0.0) | 0 (0.0) | 0 (0.0) | 0 (0.0) | 0 (0.0) | 0 (0.0) | 0 (0.0) | 0 (0.0) | 0 (0.0) | 0 (0.0) |
| Terminated by |  |  |  |  |  |  |  |  |  |  |
| sponsor |  |  |  |  |  |  |  |  |  |  |
| Study | 0 (0.0) | 0 (0.0) | 0 (0.0) | 0 (0.0) | 0 (0.0) | 0 (0.0) | 0 (0.0) | 0 (0.0) | 0 (0.0) | 0 (0.0) |
| Termination by |  |  |  |  |  |  |  |  |  |  |
| Sponsor |  |  |  |  |  |  |  |  |  |  |
| Withdrawal | 0 (0.0) | 0 (0.0) | 0 (0.0) | 0 (0.0) | 0 (0.0) | 0 (0.0) | 0 (0.0) | 1 (5.0) | 0 (0.0) | 1 (0.6) |
| by Subject |  |  |  |  |  |  |  |  |  |  |
| Other | 0 (0.0) | 0 (0.0) | 0 (0.0) | 0 (0.0) | 0 (0.0) | 0 (0.0) | 0 (0.0) | 0 (0.0) | 0 (0.0) | 0 (0.0) |

### Safety

Reactogenicity for all formulations was generally mild in severity (Figure 1) and of short duration. Both adjuvants increased the frequency of reported AEs. The frequency and severity of AEs were similar after the first and second doses in the unadjuvanted CoVLP and CoVLP+CpG1018 groups but increased after the second dose in subjects who received AS03-adjuvanted formulations. Details of solicited AEs and TEAEs by treatment group are provided in Suppl. Materials: Tables S4-S9).

**Figure 1.**
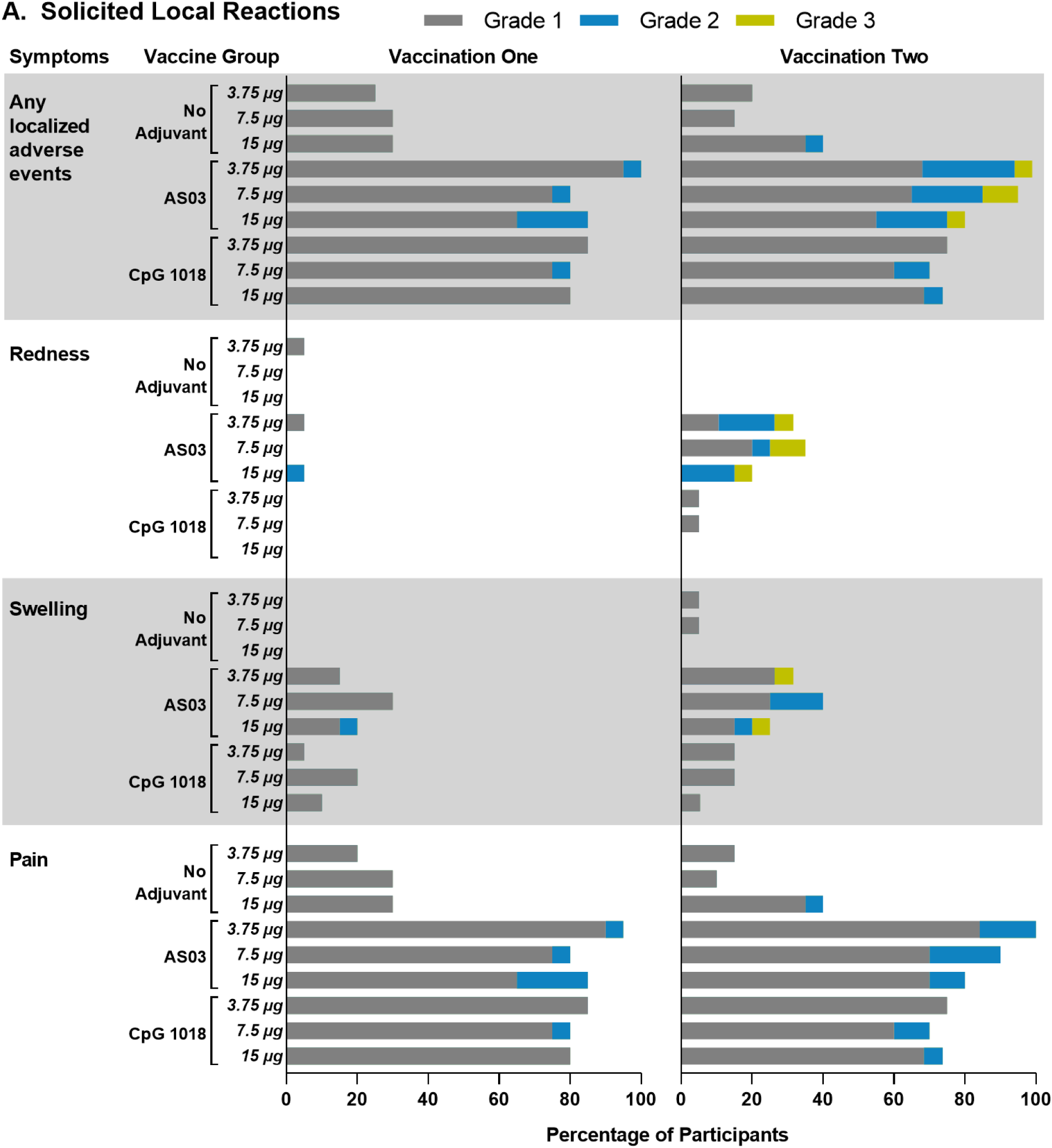
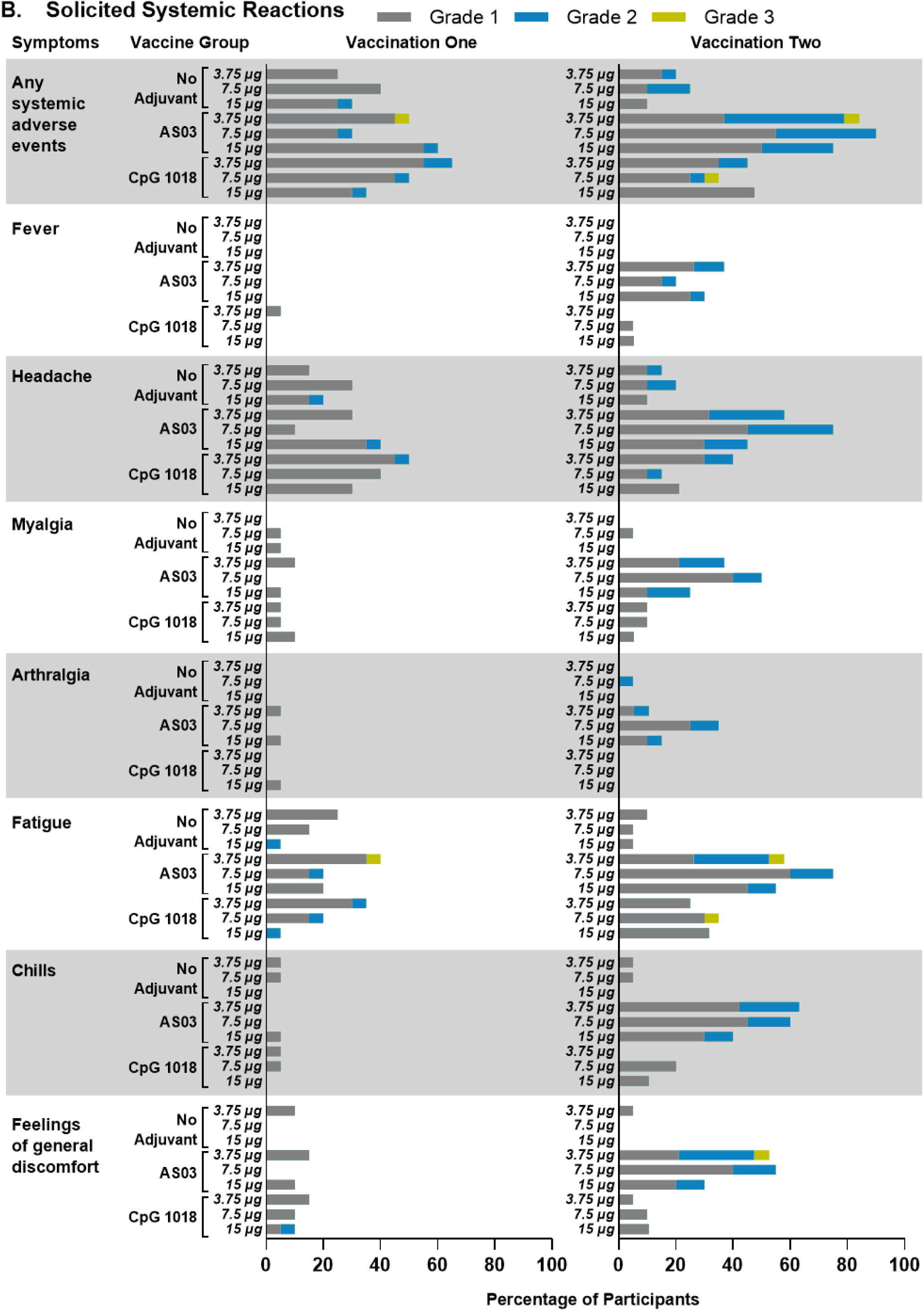
Solicited Local and Systemic AEs 7-Days After the First or Second Vaccine Dose. Subjects were monitored for solicited local (panel A) and systemic (panel B) AEs from the time of vaccination through 7 days after vaccine administration. There was no grade 4 (life-threatening) events. Participants who reported 0 events make up the remainder of the 100% calculation (not shown). If any of the solicited AEs persisted beyond Day 7 after each vaccination (when applicable), it was recorded as unsolicited AEs.

There was no consistent impact of CoVLP dose level on safety outcomes in any group. After the first dose, 74.3% of participants reported >1 solicited AE, 66.5% reporting a local reaction and 39.7% reporting >1 systemic event. Pain at the injection site was the most common local reaction (66.5%) while headache and fatigue were reported by 25.7% and 20.7% respectively. The incidence of headache and fatigue were generally higher in the adjuvanted treatment groups. AEs were mostly mild-moderate Grade 1-2) and only one Grade 3 report of fatigue that started the evening following vaccination and resolved the same day. After the second dose, 68.5% of participants reported >1 solicited AE, 62.9% reporting a local reaction and 47.8% reporting >1 systemic event. Pain at the injection site was again the most reported local reaction (61.2%) while headache and fatigue were reported by 33.1% and 33.1% respectively. Again, most symptoms were mild but there were more moderate AEs after the second dose. Nine Grade 3 solicited severe AEs (fatigue, redness at injection site, swelling at injection site, feeling of general discomfort or uneasiness) were reported in 6 subjects after the second dose. All but one of the Grade 3 reactions were reported by subjects who receivedAS03-adjuvanted formulations. One Grade 3 reaction occurred after the second dose in the CoVLP (7.5 μg)+CpG1018 group. All Grade 3 AEs resolved in one to four days. No clinically significant lab abnormalities were reported after any dose. No SAEs, AESIs or pregnancy exposures have been reported at the time of writing.

### Immunogenicity: Antibody Response

A small number of subjects (12/180; 6.7%) had detectable pre-existing antibodies for the spike protein in one or more of the assays used. As illustrated in Figure 2, unadjuvanted CoVLP elicited no detectable antibody response after the first dose and humoral responses after even the second dose were modest and inconsistent. Although a minor dose-effect for the unadjuvanted CoVLP was seen on the anti-spike IgG response (ELISA) after the second vaccination, the responses in the two NAb assays remained low and variable even at the highest dose tested. Both adjuvants had a significant impact on antibody responses at all dose levels. Although there was no convincing effect of increasing CoVLP dose for either adjuvant, there was a trend towards increasing consistency of response in the CoVLP+CpG1018 groups at the higher CoVLP doses (ie: a greater response and a larger proportion of subjects responding). Although both adjuvants elicited modest IgG titers after the first dose, only the groups that received COVLP+AS03 formulations mounted significant NAb responses at D21 (36/60; 60%) and across all dose levels (ie: overall GMT of 33.3 in the pseudovirus assay). Both adjuvants induced more robust responses after the second dose with the large majority of subjects at all dose levels mounting a ≥4-fold rise in total IgG (117/118; 99.1%) and in both NAb assays (105/112; 93.8% in the pseudovirion assay and 106/116; 91.3% in the PRNT). At all dose levels, both anti-spike IgG and pseudovirion NAb titers at D42 were 10- to 50-fold higher in subjects who had received AS03-adjuvanted formulations compared to those who had received CpG1018-adjuvanted formulations. For example, after the second CoVLP(3.75 μg) dose, the GMTs in the PRNT were 7.2 for CoVLP alone, 56.6 for CoVLP+CpG1018 and 811.3 for CoVLP+AS03. Overall, the levels of NAb induced in the groups that received two doses of CoVLP with an adjuvant were either similar to (CoVLP+CpG1018) or substantially greater (CoVLP+AS03) than those seen in subjects 3-4 weeks after recovering from natural Covid-19 infection. Details of serologic response results by treatment group are presented in Suppl. Materials: Tables S10-S12.

**Figure 2.**
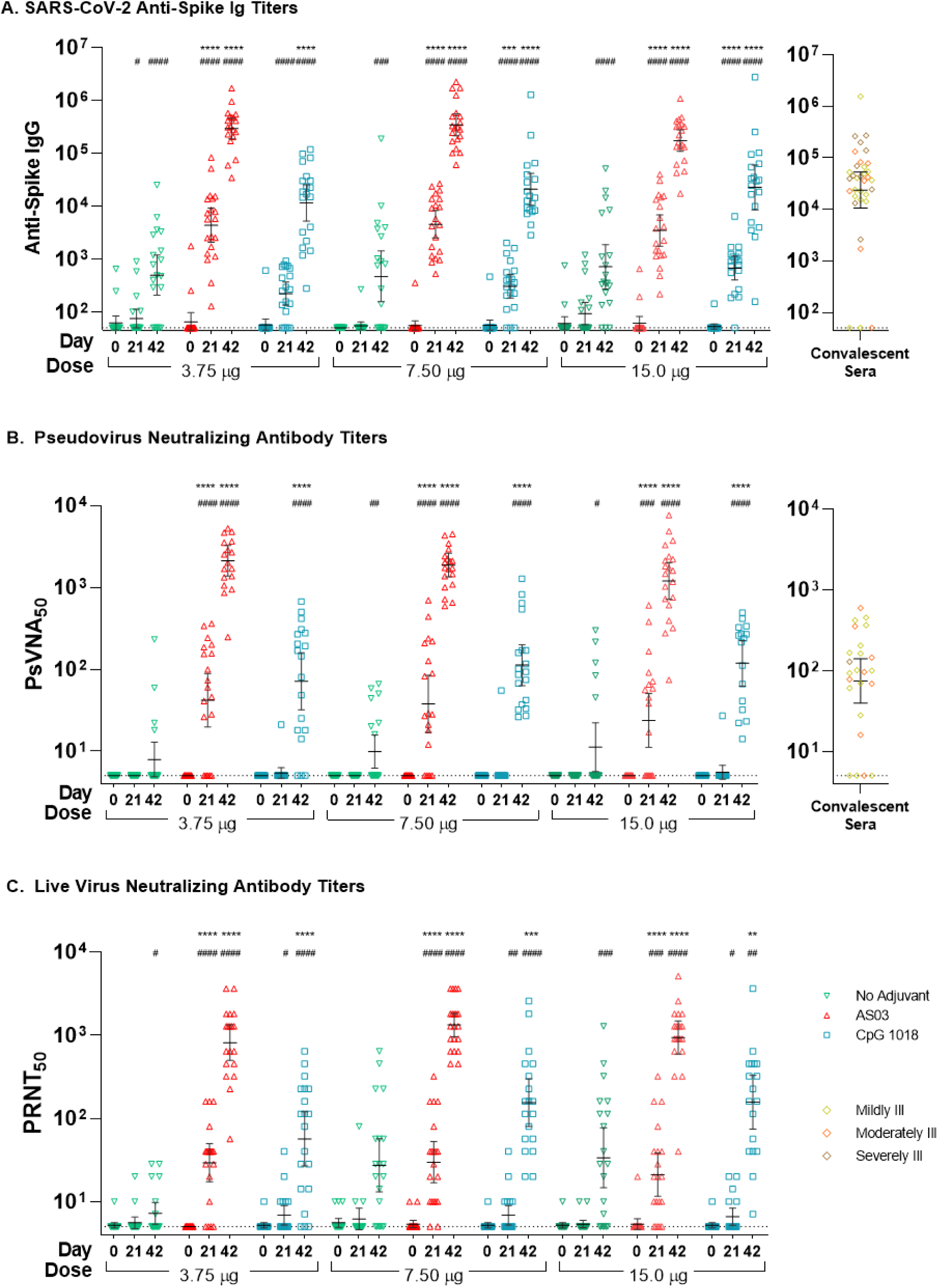
CoVLP Humoral Response. Serum antibodies of subjects vaccinated with 3.75, 7.5, or 15 µg CoVLP with or without AS03 or CpG1018 adjuvant, were measured to spike protein by ELISA (panel A) or by neutralization of pseudovirus (panel B), or live virus (panel C). Convalescent sera from recovered COVID-19 infected mildly, moderately, or severely ill patients were analyzed by anti-spike ELISA (n=35), and sera from mildly or moderately ill patients and one severely ill patient were analyzed by pseudovirion neutralization assay (n=21); results are shown in the right panels. All results are presented as reciprocal mid-point titers. Bars indicate geometric means. Error bars indicate 95% confidence intervals. Significant differences between days 0 and 21, or 0 and 42, for each vaccine regimen are indicated by # (#p<0.05, ##p<0.01, ###p<0.001, ####p<0.0001; unpaired T-test of log-transformed values, GraphPad Prism, v8.1.1). Significant differences between unadjuvanted and adjuvanted regimens for days 21 and 42 are indicated by * (*p<0.05, **p<0.01, ***p<0.001, ****p<0.0001; 2-way ANOVA of log-transformed values, GraphPad Prism, v8.1.1).

### Correlation between Neutralizing Antibody Assays

There was a strong correlation between the VSV pseudovirion neutralization assay and the PRNT for responses in all groups: r =0.85 (p < 0.0001) after the first (Figure 3A) and r = 0.68 (p < 0.0001) second vaccinations (Figure 3B).

**Figure 3.**
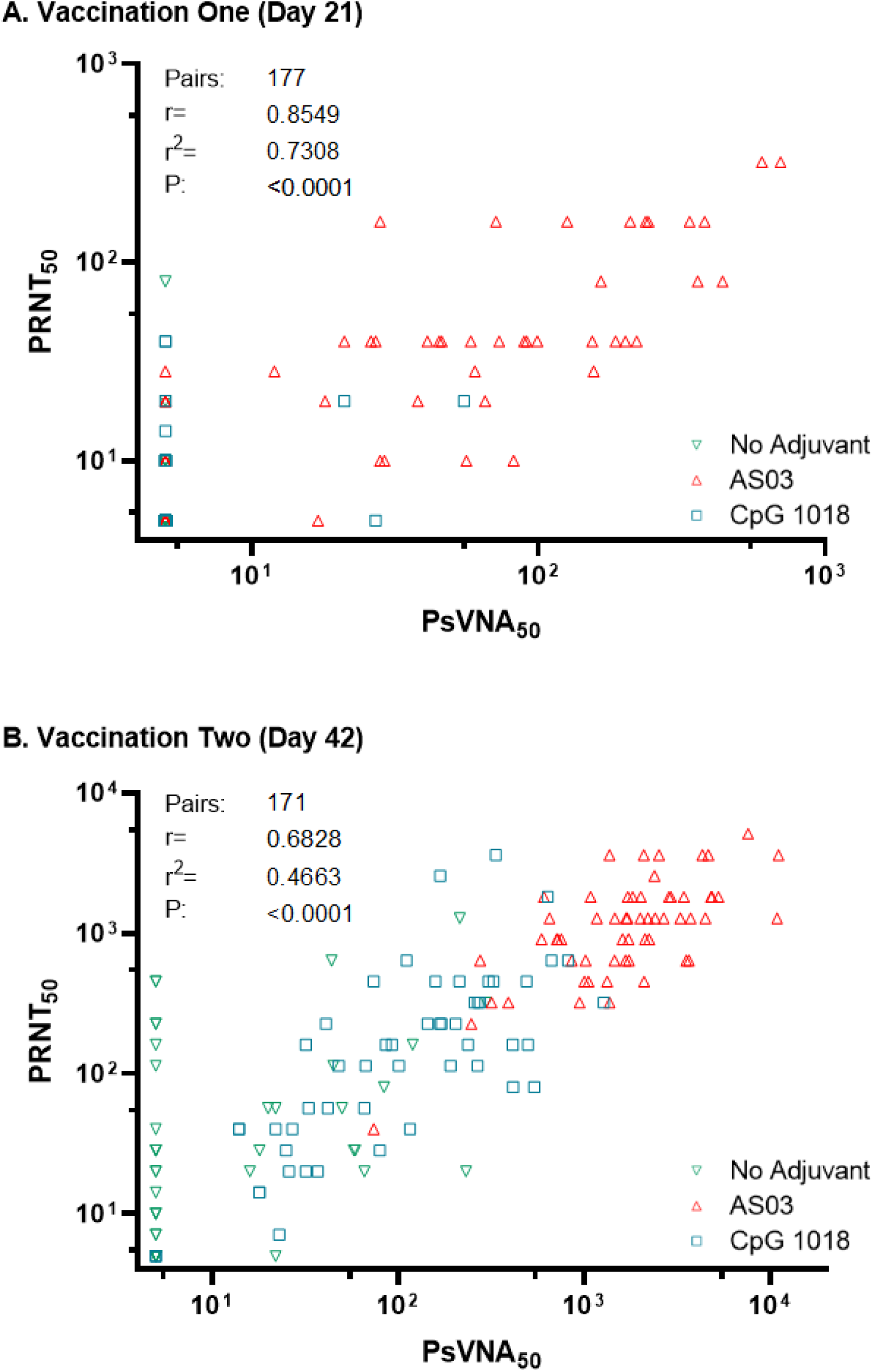
Correlation Between Assays Quantifying Neutralizing Antibody Titers. Neutralizing serum antibodies of subjects vaccinated with 3.75, 7.5, or 15 µg CoVLP with or without AS03 or CpG1018 adjuvant, were measured by neutralization of pseudovirion (x-axis) or live virus (y-axis) twenty-one days after the first vaccination (panel A) or twenty-one days afters the second vaccination (panel B). Results are presented as reciprocal titers. R are Pearson correlation coefficients (GraphPad Prism, v8.1.1).

### Immunogenicity: T Cell Response

As illustrated in Figure 4, the IFNγ and IL-4 responses in PBMC (ELISpot) elicited by CoVLP ± adjuvants were more variable than the antibody responses. A substantial minority of subjects in all groups had what appeared to be pre-existing IFNγ responses to the S protein peptide pool that were, in some subjects, substantial (ie: >200 spots)^33^. Although low level ‘background’ IL-4 activity was seen in a small number of subjects, these responses were close to the limit of detection of the assay used (generally <10 spots). Unlike antibody responses, CoVLP alone was able to induce a substantial IFNγ response and, to a lesser extent, IL-4 response in many subjects after the second doses at all dose levels but that was most consistently seen at the highest dose (CoVLP 15μg). Both adjuvants generally increased IFNγ and IL-4 responses above background levels after the first dose that were further and substantially increased in magnitude and consistency by the second dose. Compared to unadjuvanted CoVLP, IFNγ responses were slightly higher and IL-4 responses were slightly lower in the CpG1018-adjuvanted groups but most of these differences did not reach statistical significance. Once again, the IFNγ and IL-4 responses to the CoVLP+AS03 formulations at all dose levels were 10 to 50-fold higher than those seen in the equivalent CoVLP+CpG1018 groups. For example, at the 3.75 µg dose level: median IFNγ and IL-4 responses at D42 were 628 and 445 in the CoVLP+AS03 group and 49 and 4 in the CoVLP+CpG1018 group. Details of cellular response results by treatment group are provided in Suppl. Materials: Tables S13 - S14.

**Figure 4.**
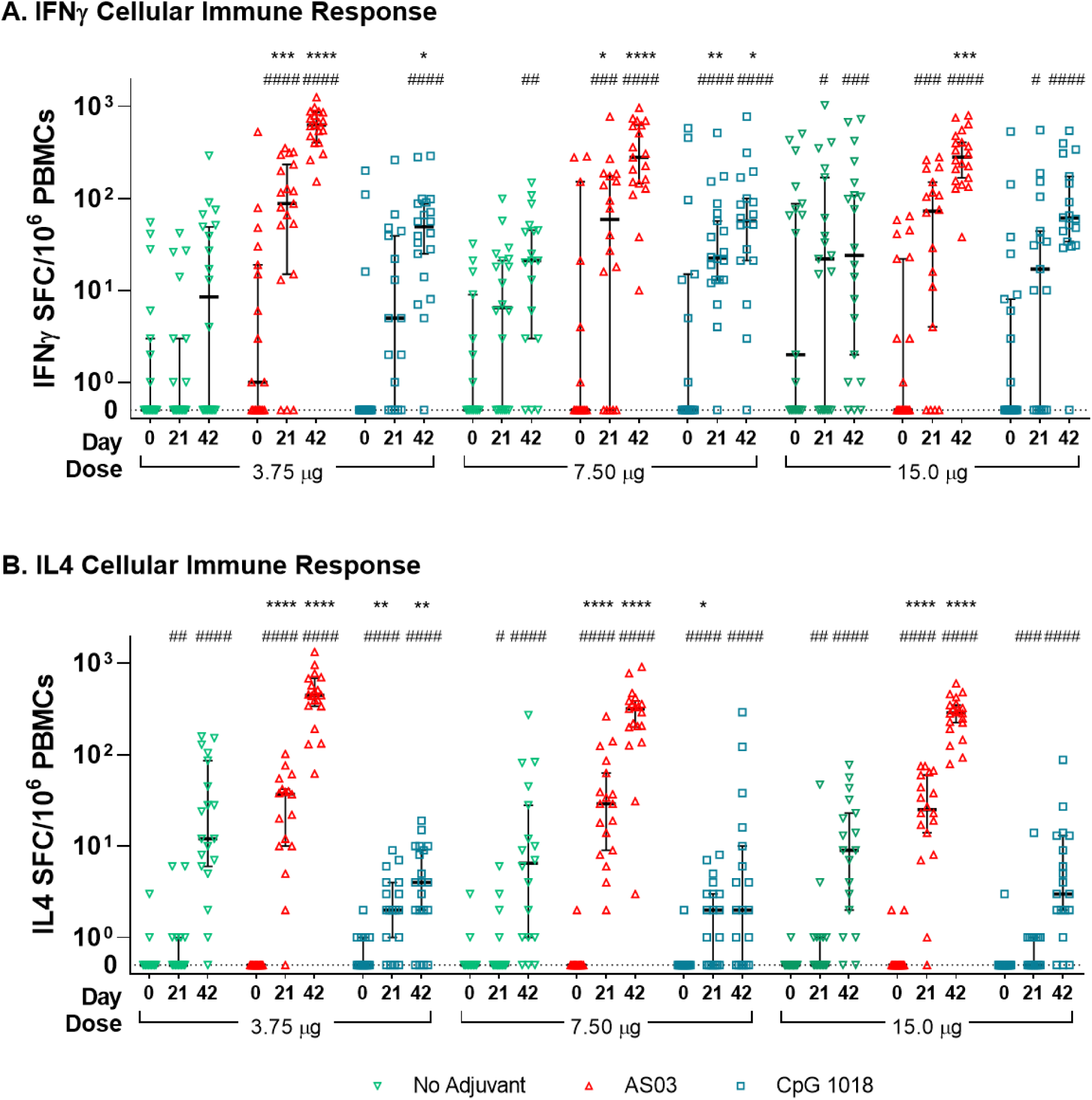
IFN-γ and IL-4 secreting cells as measured by ELISpot at Day 0, 21, and 42. Frequencies of antigen-specific T-cells producing interferon-gamma (Panel A) and interleukin-4 (Panel B) cellular immune responses at baseline (day 0) and 21 days after one immunization (day 21) or two immunizations (day 42) with 3.75, 7.5 or 15 µg doses of CoVLP with or without adjuvants (CpG1018 and AS03) after restimulation ex vivo with recombinant spike peptide pool. Bars indicate medians and error bars indicate 95% CI. Significant differences between days 0 and 21 or between day 0 and 42 for each vaccine regimen are indicated by # (^#^P<0.05, ^##^P<0.01, ^###^P<0.001, ^####^P<0.0001; Unpaired T-test. Comparisons for each test to day 0 data are to the same pre-vaccination data set that include all subjects. The figure illustrates matched subject data. GraphPad Prism, v8.1.1). Significant difference between adjuvanted vaccine and unadjuvanted vaccine regiments at day 21 and 42 are indicated by * (*P<0.05, **P<0.01, ***P<0.001, ****P<0.0001; Kruskal-Wallis, GraphPad Prism, v8.1.1).

## Discussion

This phase 1 study was designed to select the CoVLP formulation (ie: dose level ± adjuvant) and the number of doses needed to generate a consistent immune response in healthy adults with an acceptable safety profile. Although unadjuvanted CoVLP had the lowest reactogenicity and was recognized by the immune system after the second dose, the immune responses measured (i.e.:anti-spike IgG, NAb, IFNγ and IL-4 ELISpots) were generally modest. These observations are consistent with studies of plant-derived VLP vaccines bearing HA trimers of avian influenza strains ^30,34^ in which serologic and T cell responses were observed with unadjuvanted formulations at doses >15 µg but were seen at lower doses when administered with an oil-in-water adjuvant (GLA-SE)^34^. Although we observed no dose effect with unadjuvanted CoVLP for antibody or cellular responses, it is possible that higher doses of unadjuvanted CoVLP would have induced stronger responses. However, two different adjuvants were incorporated into this study because of their potential to induce more robust responses at a lower CoVLP dose (ie: dose-sparing).

While sharing some characteristics (ie: documented immune enhancement, large safety databases, previously licensed)^35-38^, the inclusion of both CpG1018 and AS03 permitted us to ask multiple questions about CoVLP performance simultaneously. In the case of CpG1018, by targeting the innate toll-like receptor 9 we hoped that both arms of the adaptive immune system would be engaged with induction of humoral and cellular responses ^39^. Although the mechanism of action of oil-in-water adjuvants like AS03 is still under active investigation^40^, the combination of squalene and tocopherol (vitamin E) in AS03 has also been shown to enhance both humoral and cellular responses with several different vaccine antigens ^41,42^.

In the current study, the adjuvants performed as expected with enhancement of humoral and cellular responses to the S protein and in promoting responses at lower CoVLP doses. In both dose-sparing and enhancing immune responses, AS03 appeared to be more effective than CpG1018. For example, a substantial minority of the subjects who received even the lowest dose of CoVLP+AS03 (3.75 µg) mounted detectable NAb responses after the first dose but no similar response was seen in the CpG1018-adjuvanted groups. Furthermore, the anti-spike IgG and NAb responses were consistently higher at all dose levels in the AS03-adjuvanted groups compared to the CpG1018-adjuvanted groups although differences did not reach statistical significance at higher doses of CoVLP. The humoral response to the AS03-adjuvanted formulations demonstrated no CoVLP dose effect, particularly after the second dose. Overall, the differences between unadjuvanted and adjuvanted formulations and between the two adjuvants were less pronounced for Th1-type cell-mediated responses than for antibody responses. Nonetheless, IFNγ responses to adjuvanted formulations were consistently higher at most dose levels than responses to CoVLP alone, particularly after the second dose. Overall, IFNγ responses were strongest in the AS03-adjuvanted groups, particularly at lower CoVLP dose levels. Of course, it is possible that the IFNγ responses observed were mediated in part by other cells (eg: NK or CD8 T cells), since the ELISpot assay captures cell-mediated responses indiscriminately^44^. Additional flow cytometry evaluations are warranted to characterize the phenotype of these cellular responses more comprehensively and in particular, to better understand the relative contribution of CD4 T cells to the observed responses. The IL-4 response was also consistently higher in the AS03-adjuvanted groups compared to the unadjuvanted or CpG1018-adjuvanted groups. In the AS)3-adjuvanted groups, the IL4 response was 10-fold lower than the IFNγ responses after the first dose but rose to near equivalence with the second dose suggesting a balanced Th1- and Th2-type response.

It is noteworthy that a substantial proportion of the subjects in this study appeared to have pre-existing IFNγ responses to the S protein peptide pool used for PBMC restimulation. Such cross-reactive T cell memory, possibly due to prior exposure to common human coronaviruses has been seen in 40-60% of adults and may provide some protection against highly pathogenic strains ^33^. Prior exposure to circulating coronaviruses may also explain the small number of subjects in this study (6.7%) who were ‘seronegative’ at screening based upon a commercial N-based ELISA but who were sero-positive at D0 in our assays targeting the S protein.

Together, these immune outcome data suggest that CoVLP alone can elicit both humoral and cellular responses that are Th1-biased to some extent and that this pattern of response is reinforced when adjuvanted with CpG1018. In contrast, when CoVLP is adjuvanted with AS03, the response was faster and more balanced with evidence of both Th1- and Th2-type activation. Although Th2-deviated responses have been implicated in VED (e.g.: early RSV vaccine)^45^ and are a theoretical risk for vaccines targeting pathogenic coronaviruses ^46^, the engagement of T cells is also critical for B-cell maturation and the induction of strong and durable antibody responses ^47,48^.

The safety profile of CoVLP alone was relatively benign at all dose levels but, as expected, both the frequency and the intensity of local and systemic AEs was increased with either adjuvant. Compared to the first dose, the frequency of Grade 2 or Grade 3 AEs also increased after the second dose in all groups that received adjuvanted formulations. All AEs reported, regardless of dose level, adjuvant or initial intensity, were transient and resolved rapidly. Although one subject did not receive a second dose of vaccine due to a grade 3 AE (per Protocol), no subject voluntarily withdrew from the study due to any AE/SAE. Overall, the reactogenicity of the adjuvanted CoVLP formulations was similar to that reported for other candidate SARS-CoV-2 vaccines in early-phase clinical studies. Although hypersensitivity to plant material is a theoretical risk with any plant-derived product, no subject in the current study had an allergic-type reaction, consistent with Medicago’s safety database of >14,000 subjects who have received one or two doses of plant-derived influenza vaccine candidates^49^.

Like any early-phase clinical trial, this study has several limitations beyond the obvious concern regarding small group size when testing multiple dose levels and formulations (n=20/group). For comparisons between formulations however, the small group risk was mitigated by the fact that results across the three CoVLP dose levels were highly consistent (n=60 for CoVLP alone or with each adjuvant). Another obvious concern is the relatively limited range of immune response parameters available at the time of writing. Although the data presented herein provide a ‘first look’ at the immune response induced by the different formulations, the immune profiles will be further investigated in the coming months. Finally, like all other early-phase trials, our study assessed only the short-term (day 42) response to vaccination and the durability of these responses will only become apparent as subjects are followed for longer periods of time. However, based on the robust high antibody and balanced cellular responses to the adjuvanted CoVLP formulations and our previous experience with plant-derived VLP vaccines targeting influenza, we anticipate that the CoVLP-induced responses are likely to last for at least 6 months^30, 50^.

In conclusion, this first trial of CoVLP alone or adjuvanted with either CpG1018 or AS03 suggests that our plant-derived candidate vaccine is well-tolerated and immunogenic. Its immunogenicity, particularly at low doses, is dramatically enhanced by the presence of an adjuvant. Based upon the totality of non-clinical and clinical results available and the obvious advantages of dose-sparing in a pandemic, a two-dose schedule of CoVLP at 3.75 μg/dose adjuvanted with AS03 will be carried forward into further studies planned for Canada, the USA and other countries.

## Data Availability

This is an interim analysis at Day 42 of a Phase 1 trial. Complete Data will be made available upon conmpletion of the safety follow-up period only and upon Request.

## Appendix

### Author Contributions

All authors contributed significantly to the research work. All company authors had full access to the Tables and Figure data. BJW and NL made the final decision to submit the manuscript.

## Acknowledgements

The authors would like to acknowledge F. Roman, P. Boutet, Robbert Van Der Most, M. Ceregido Perez, M. Koutsoukos and G. Preyon from GSK as well as R. Spencer, R. Janssen, D. Campbell and D. Novack from Dynavax for critical review of the manuscript. The authors also wish to acknowledge all the Medicago employees, their contractors and all of the volunteers who participated in the study as well as the site investigators and their staff who conducted the studies with a high degree of professionalism.

## Supplementary Appendix to

## Supplementary Methods

### Vaccination Stopping Rules

Safety monitoring of safety signals were and will be performed throughout the study. Stopping rules or conditions for stopping this clinical trial would occur if there was clear evidence of harm or harmful effects such as SAEs related to the treatment (study vaccine). A SAE which was thought to be unrelated to the study vaccine would not warrant stopping the trial.

The following event(s) may result in a halt to the study, for further review and assessment of the event(s):

- Any death occurring during the study;
- Any vaccine-related SAE during the study;
- Any life-threatening (Grade 4) vaccine-related AE during the study;
- If 10 % or more of subjects in a single treatment group, experience the same or similar listed event(s) that cannot be clearly attributed to another cause:
  - a severe (Grade 3 or higher) vaccine-related AE during the study;
  - a severe (Grade 3 or higher) vaccine-related vital sign(s) abnormality;
  - a severe (Grade 3 or higher) vaccine-related clinical laboratory abnormality.

In the case that a pre-defined safety signal is met in any treatment group, subsequent dosing will result in at least a transient halt in the study to permit a complete evaluation of the reported event(s) and to consult an IDMC. A decision as to whether the study can progress as planned must be made and documented in the event of any safety signal. If a stopping rule has been met once all subjects have been vaccinated in the study, the IDMC will be notified by a Note To File (NTF) for their information purposes.

### Immunological Assay Method Details

#### Evaluation of SARS-CoV-2 Humoral Response

#### SARS-CoV-2 Spike Protein ELISA

Briefly, SARS-Cov2 Spike protein (SARS-Cov2/Wuhan/2019, Immune Technology Corp.) was coated onto a flat-bottom 96-well microplate at a concentration of 1ug/mL in sodium carbonate 50mM (overnight; 4°C). Following washing steps (PBS-Tween), plates were blocked using Blotto 5% (Rockland Inc.) in PBS (1-2 hours; 37°C). Following washing steps, serially diluted sera (starting dilution 1/100, 4-fold dilutions, 8 dilutions, in PBS-Tween-Blotto) were added to the wells, in duplicates, and incubated at 37°C for 1 hour. Plates were washed and incubated with secondary antibody (anti-Human IgG (H+L) antibody, Peroxidase-labeled, Seracare), diluted at 1/20 000 in PBS-Tween-Blotto and incubated at 37°C for 1 hour. Plates were washed and incubated with peroxidase substrate (SureBlue TMB, Seracare) for 20 minutes at room temperature. Reaction was stopped using hydrochloric acid and absorbance was read at 450nm, within 2 hours (Variaskan Flash microplate reader, Thermo Scientific). Optical density (OD) results for the serial dilutions were used to generate a 4-parameter logistic regression (4PL). The titer was defined as the reciprocal dilution of the sample for which the OD is equal to a fixed cut-point value. Samples below cut-point were attributed a value of 50 (half the minimum required dilution).

#### SARS-CoV-2 Pseudovirus Neutralisation Assay (PNA)

Neutralizing antibody analysis was performed using a cell-based pseudotyped virus neutralisation assay (Nexelis, Quebec, Canada). Pseudotyped virus particles were first generated using a genetically modified Vesicular Stomatitis Virus backbone from which the glycoprotein G was removed and luciferase reporter introduced (rVSVΔG-luciferase, Kerafast) to allow quantification using relative luminescence units (RLU). This rVSVΔG–Luc virus was transduced into HEK293T cells that had previously been transduced with SARS-Cov-2 Spike glycoprotein (NXL137-1 in POG2 containing 2019-nCOV Wuhan-Hu-1; Genebank: MN908947) from which the last nineteen amino acids of the cytoplasmic tail were removed (rVSVΔG-Luc-Spike ΔCT).

Serial dilutions (starting dilution of 1/10; 2-fold; 8 dilutions, in complete growth media) of the heat-inactivated human sera (56°C; 30min) were prepared in a 96-well plate, in duplicates. The SARS-Cov-2 pseudovirus (in complete growth media) was added to the sera dilutions to reach a target concentration equivalent to approximately 150,000 RLU/well and mixture was incubated at 37°C with 5% CO_2_ supplementation for 1 hour. Serum-pseudovirus complexes were then transferred onto plates previously seeded overnight with Vero E6 cells (ATCC CRL-1586), expressing ACE-2 receptor, and incubated at 37°C with 5% CO_2_ supplementation for 20-24 hours. Once incubation was completed, cells were lysed and samples equilibrated using ONE-Glo EX luciferase assay system (Promega), incubated for 3 minutes at room temperature, and luminescence level read using a luminescence plate reader (i3x plate reader, Molecular Devices). The resulting RLU was inversely proportional to the level of neutralizing antibodies present in the serum. For each sample, the neutralizing titer was established as the reciprocal dilution corresponding to the 50% neutralization (NT50), when compared to the pseudoparticle control. The NT50 was interpolated from a linear regression using the two dilutions flanking the 50% neutralisation. Samples below cut-off were attributed a value of 5 (half the minimum required dilution).

#### SARS-CoV-2 Microneutralization CPE-based assay

Neutralizing antibody analysis was also performed using a cell-based cytopathic effect (CPE) assay (VisMederi, Sienna, Italy). Sera sample were first heat inactivated (56°C; 30min) and then serially diluted (starting dilution of 1/10, 2-fold; 8 dilutions, in complete growth media). Wild-type SARS-Co-2 virus (2019 nCOV ITALY/INMI1, provided by EVAg; Genebank: MT066156) was then added at final concentration of 25 TCID50/mL (in complete growth media), and plates were incubated for 1 hour at 37°C with 5% CO_2_ supplementation. At the end of the incubation, the mixture was transferred onto duplicate 96-well microtiter plates pre-seeded overnight with Vero E6 cells (ATCC CRL-1586), expressing ACE-2 receptor. Plates were then incubated for 3 days at 37°C with 5% CO_2_ supplementation. Cytopathic effect (CPE) was then quantified using an inverted optical microscope. The microneutralization titer (MNt) was defined as the reciprocal of the highest sample dilution that protects from CPE at least 50% of the cells. If no neutralization was observed, samples were attributed a titer value of 5 (half the minimum required dilution).

#### Evaluation of SARS-CoV-2 Cell-Mediated Immune Response

##### Interferon-γ ELISpot

Cell-mediated immune response was evaluated using an Interferon-γ ELISpot assay (Human IFN-γ ELISpot assay, Cellular Technology Limited (CTL), USA). Cryopreserved peripheral blood mononuclear cells (PBMC) were rapidly thawed and allowed to rest between 2 to 3 hours at 37°C with 5% CO_2_ supplementation, in CTL-Test media supplemented with 1% Glutamine and 1% Penicillin/Streptomycin. Cells were enumerated and dispensed at 0.5×10^6^ cells per well, in duplicates, onto PVDF filter plates pre-coated with an IFN-γ specific capture antibody. Cells were stimulated using a pool of peptides (15-mer peptides) overlapping the full sequence of SARS-Cov-2 spike protein (USA-CA2/2020, Genbank: MN994468.1) at a concentration of 2.19 µg/mL, for 18-24 hours, at 37°C with 5% CO_2_ supplementation. After washes (PBS-Tween), biotinylated anti-interferon-γ detection antibody was added to the plates and incubated for 2 hours at room temperature after which, following another round of washes, a streptavidin-alkaline phosphatase conjugate was added and incubated for 30 minutes at room temperature. After washing steps, substrate solution was added, incubated at room temperature for 15 minutes, after which plate was rinsed and left to air-dry pending spot enumeration, using an ELISpot reader (ImmunoSpot S6 Universal Analyzer, Cellular Technology Limited). Mean of peptide pool stimulation duplicates was calculated and normalized using the mean of the negative control replicates (control media) and multiplied by a factor of 2 to express cells counts per million cells.

#### Interleukin-4 (IL-4) ELISpot

Cell-mediated immune response was evaluated using an IL-4 ELISpot assay (Human IL-4 ELISpot assay, Cellular Technology Limited (CTL), Cleveland, OH, USA). Cryopreserved peripheral blood mononuclear cells (PBMC) were rapidly thawed and allowed to rest between 2 to 3 hours at 37°C with 5% CO_2_ supplementation, in CTL-Test media supplemented with 1% Glutamine and 1% Penicillin/Streptomycin. Cells were enumerated and dispensed at 0.5×10^6^ cells per well, in duplicates, onto PVDF filter plates pre-coated with an IL-4 specific capture antibody. Cells were stimulated using a pool of peptides (15-mer peptides) overlapping the full sequence of SARS-Cov-2 spike protein (USA-CA2/2020, Genbank: MN994468.1) at a concentration of 2.19 µg/mL, for 32-48 hours, at 37°C with 5% CO_2_ supplementation. After washes (PBS-Tween), biotinylated anti-IL-4 detection antibody was added to the plates and incubated for 2 hours at room temperature after which, following another round of washes, a streptavidin-alkaline phosphatase conjugate was added and incubated for 30 minutes at room temperature. After washing steps, substrate solution was added, incubated at room temperature for 15 minutes, after which plate was rinsed and left to air-dry pending spot enumeration, using an ELISpot reader (ImmunoSpot S6 Universal Analyzer, Cellular Technology Limited). Mean of peptide pool stimulation duplicates was calculated and normalized using the mean of the negative control replicates (control media) and multiplied by a factor of 2 to express cells counts per million cells.

### Convalescent sera description

Sera from Covid-19 convalescent patients were collected from a total of 34 individuals with confirmed disease diagnosis. Time between the onset of the symptoms and sample collection varied between 27 and 105 days. Three samples were collected by Solomon Park (Burien, WA, USA), 20 by Sanguine BioSciences (Sherman Oaks, CA, USA) and 11 from McGill University Health Centre. Disease severity were ranked as mild (Covid-19 symptoms without shortness of breath), moderate (shortness of breath reported), and severe (hospitalized). These samples were analysed in parallel of clinical study samples, using the same assay as described above. Samples from severely ill were analyzed by serum IgG ELISA only. Demographic characteristics are provided in Supplementary Table 1.

**Supplementary Table 1.** Summary of Covid-19 Patients Providing Convalescent Sera.

| Characteristics | Mild | Moderate | Severe |
| --- | --- | --- | --- |
| Subjects | 16 | 8 | 11 |
| Sex, n (%) |  |  |  |
| Male | 10 (62.5%) | 2 (25.0%) | 8 (72.7%) |
| Female | 5 (31.3%) | 6 (75.0%) | 3 (27.3%) |
| Race, n (%) |  |  |  |
| White | 7 (43.8%) | 6 (75.04%) | 5 (45.5%) |
| Black or African American | 1 (6.3%) | 0 (0.0%) | 5 (45.5%) |
| Asian | 4 (25.0%) | 2 (25.0%) | 0 (0.0%) |
| Ethnicity, n (%) |  |  |  |
| Hispanic/Latinx | 2 (12.5%) | 1 (12.5%) | 1 (9.1%) |
| Age: |  |  |  |
| Mean ± SD | 42.7 ± 13.6 | 37.8 ± 13.0 | 51.9 ± 16.0 |
| Median (range) | 39 (20-66) | 40.5 (19-58) | 50.0 (28-82) |

SD: Standard Deviation

### Adverse Event Severity Evaluation and the Monitoring of Adverse Events of Special Interest

#### Adverse Event of Special Interest (AESI)

##### AESI for CoVLP Vaccine – Vaccine Enhanced Disease (VED)

Safety signal of VED after exposure to the Coronavirus-Like Particle COVID-19 Vaccine was closely monitored and assessed by retrieving data for this AESI as follows: AEs within the system organ class (SOC): immune system disorders and high level group term (HLGT): lower respiratory tract disorders (excluding obstruction and infection), cardiac disorders, signs and symptoms not elsewhere classified (NEC), vascular disorders, heart failures NEC, arteriosclerosis, stenosis, vascular insufficiency and necrosis, cardiac arrhythmias, myocardial disorders, and vascular hemorrhagic disorders. High level term (HLT): renal failure and impairment and preferred term (PT): pericarditis, coagulopathy, deep vein thrombosis, pulmonary embolism, cerebrovascular accidents, peripheral ischemia, liver injury, Guillain-Barre syndrome, anosmia, ageusia, encephalitis, chilblains, vasculitis, erythema multiforme (based on standardized MedDRA^®^ classification)^1, 2^ that require inpatient hospitalization (≥ 24 hours) and have laboratory confirmed SARS-Cov-2 infection will be monitored for assessment of any potential case of VED.

##### AESI for CoVLP Vaccine – Hypersensitivity Reactions

All reported events were also monitored for hypersensitivity reactions after exposure to the Coronavirus-Like Particle COVID-19 Vaccine. In eight clinical studies conducted to date with the Quadrivalent VLP Influenza Vaccine (QVLP) produced using similar plant-based technology, all reported events were monitored for a possible hypersensitivity component (events were searched using both narrow and broad standardized MedDRA^®^ queries). Based on these data, there was a single case of possible early anaphylactic reaction associated with use of QVLP in humans. A small number of subjects had potential hypersensitivity reactions judged to be related to vaccine administration (no more than 0.3 % of subjects in any given QVLP treatment group experienced one of these events) and the events were distributed fairly evenly among treatment groups, including the placebo and the active comparator groups. However, since severe reactions are considered to be an important potential risk (based on the theoretical risk that using plants for the production of biotherapeutics may induce hypersensitivity), Medicago required that appropriate medical treatment and supervision were available to manage any possible anaphylactic reactions in this study. To collect data on these events, Medicago closely monitored and assessed allergic reactions assessed by the site Investigators as related to the Investigational product as AESIs.

##### AESI for Adjuvant – Potential Immune-Mediated Diseases (pIMD)

Potential immune-mediated diseases are a subset of AEs that include autoimmune diseases and other inflammatory and/or neurologic disorders of interest which may or may not have an autoimmune aetiology.

**Supplementary Table 2.** List of Potential Immune-Mediated Diseases.

| **Neuroinflammatory disorders** | **Musculoskeletal disorders** | **Skin disorders** |
| --- | --- | --- |
| • Cranial nerve neuropathy, including paralysis and paresis (eg, Bell's palsy).<br>• Optic neuritis.<br>• Multiple sclerosis.<br>• Transverse myelitis.<br>• Guillain-Barré syndrome, including Miller Fisher syndrome and other variants.<br>• Acute disseminated encephalomyelitis, including site specific variants, eg, noninfectious encephalitis, encephalomyelitis, myelitis, myeloradiculoneuritis.<br>• Myasthenia gravis, including Lambert-Eaton myasthenic syndrome.<br>• Demyelinating peripheral neuropathies and plexopathies including:<br>- Chronic inflammatory demyelinating polyneuropathy.<br>- Multifocal motor neuropathy.<br>- Polyneuropathies associated with monoclonal gammopathy.<br>• Narcolepsy.<br>• Tolosa Hunt syndrome. | • Systemic lupus erythematosus and associated conditions.<br>• Systemic scleroderma (systemic sclerosis), including:<br>- Diffuse scleroderma.<br>- CREST syndrome.<br>• Idiopathic inflammatory myopathies, including:<br>- Dermatomyositis.<br>- Polymyositis.<br>• Anti-synthetase syndrome.<br>• Rheumatoid arthritis and associated conditions including:<br>- Juvenile idiopathic arthritis.<br>- Still's disease.<br>• Polymyalgia rheumatica.<br>• Spondyloarthropathies, including:<br>- Ankylosing spondylitis.<br>- Reactive arthritis (Reiter's syndrome).<br>- Undifferentiated spondyloarthritis.<br>- Psoriatic arthritis.<br>- Enteropathic arthritis.<br>• Relapsing polychondritis.<br>• Mixed connective tissue disorder.<br>• Gout. | • Psoriasis.<br>• Vitiligo.<br>• Erythema nodosum.<br>• Autoimmune bullous skin diseases (including pemphigus, pemphigoid, and dermatitis herpetiformis).<br>• Lichen planus.<br>• Sweet's syndrome.<br>• Localized scleroderma (morphea).<br>• Cutaneous lupus erythematosus.<br>• Rosacea. |
| **Vasculitis** | **Blood disorders** | **Others** |
| • Large vessels vasculitis including:<br>- Giant cell arteritis (temporal arteritis).<br>- Takayasu's arteritis.<br>• Medium sized and/or small vessels vasculitis including:<br>- Polyarteritis nodosa.<br>- Kawasaki's disease.<br>- Microscopic polyangiitis.<br>- Wegener's granulomatosis (granulomatosis with polyangiitis). | • Autoimmune hemolytic anemia.<br>• Autoimmune thrombocytopenia.<br>• Antiphospholipid syndrome.<br>• Pernicious anemia.<br>• Autoimmune aplastic anemia.<br>• Autoimmune neutropenia.<br>• Autoimmune pancytopenia. | • Autoimmune glomerulonephritis including:<br>- IgA nephropathy.<br>- Glomerulonephritis rapidly progressive.<br>- Membranous glomerulonephritis.<br>- Membranoproliferative glomerulonephritis.<br>- Mesangioproliferative glomerulonephritis.<br>- Tubulointerstitial nephritis and uveitis syndrome. |

| **Vasculitis (continued)** | **Blood disorders (continued)** | **Others (continued)** |
| --- | --- | --- |
| - Churg–Strauss syndrome (allergic granulomatous angiitis or eosinophilic granulomatosis with polyangiitis).<br>- Buerger’s disease (thromboangiitis obliterans).<br>- Necrotizing vasculitis (cutaneous or systemic).<br>- Antineutrophil cytoplasmic antibody (ANCA) positive vasculitis (type unspecified).<br>- Henoch-Schonlein purpura (IgA vasculitis).<br>- Behcet’s syndrome.<br>- Leukocytoclastic vasculitis. |  | • Ocular autoimmune diseases including:<br>- Autoimmune uveitis.<br>- Autoimmune retinitis.<br>• Autoimmune myocarditis/cardiomyopathy.<br>• Sarcoidosis.<br>• Stevens-Johnson syndrome.<br>• Sjögren’s syndrome.<br>• Alopecia areata.<br>• Idiopathic pulmonary fibrosis.<br>• Goodpasture syndrome.<br>• Raynaud’s phenomenon. |
| **Liver disorders** | **Gastrointestinal disorders** | **Endocrine disorders** |
| • Autoimmune hepatitis.<br>• Primary biliary cirrhosis.<br>• Primary sclerosing cholangitis.<br>• Autoimmune cholangitis. | • Inflammatory bowel disease, including:<br>- Crohn’s disease.<br>- Ulcerative colitis.<br>- Microscopic colitis.<br>- Ulcerative proctitis.<br>• Celiac disease.<br>• Autoimmune pancreatitis. | • Autoimmune thyroiditis (including Hashimoto thyroiditis).<br>• Grave’s or Basedow’s disease.<br>• Diabetes mellitus type 1.<br>• Addison’s disease.<br>• Polyglandular autoimmune syndrome.<br>• Autoimmune hypophysitis. |

#### Severity of Grades for Solicited Local and Systemic Adverse Events

**Supplementary Table 3.** List of Potential Immune-Mediated Diseases.

| Symptoms | Severity |  |  |  |  |
| --- | --- | --- | --- | --- | --- |
|  | None | Grade 1<br>(Mild) | Grade 2<br>(Moderate) | Grade 3<br>(Severe) | Grade 4<br>(Potentially life-threatening) |
| **Injection Site Adverse Events (Local Adverse Events)** |  |  |  |  |  |
| **Erythema (redness)** | < 25 mm | 25 - 50 mm | 51 - 100 mm | > 100 mm | Necrosis or exfoliative dermatitis |
| **Swelling** | < 25 mm | 25 - 50 mm and does not interfere with activity | 51 – 100 mm or interferes with activity | > 100 mm or prevents daily activity | Necrosis |
| **Pain** | None | Does not interfere with activity | Repeated use of non-narcotic pain reliever for more than 24 hours or interferes with activity | Any use of narcotic pain reliever or prevents daily activity | Results in a visit to emergency room (ER) or hospitalization |
| **Solicited Systemic Adverse Events** |  |  |  |  |  |
| **Fever (°C or °F)** | < 38.0 °C<br>< 100.4 °F | 38.0 - 38.4 °C<br>100.4 - 101.1 °F | 38.5 - 38.9 °C<br>101.2 - 102.0 °F | 39.0 - 40.0 °C<br>102.1 - 104.0 °F | > 40.0 °C<br>> 104.0 °F |
| **Headache** | None | No interference with activity | Repeated use of non-narcotic pain reliever for more than 24 hours or some interference with activity | Significant; any use of narcotic pain reliever or prevents daily activity | Results in a visit to emergency room (ER) or hospitalization |
| **Fatigue** | None | No interference with activity | Some interference with activity | Significant; prevents daily activity | Results in a visit to emergency room (ER) or hospitalization |
| **Muscle aches** | None | No interference with activity | Some interference with activity | Significant; prevents daily activity | Results in a visit to emergency room (ER) or hospitalization |
| **Joint aches, chills, feeling of general discomfort or uneasiness (malaise), swelling in the axilla, swelling in the neck, swelling in the groin, swelling in the chest wall** | None | No interference with activity | Some interference with activity not requiring medical intervention | Prevents daily activity and requires medical intervention | Results in a visit to emergency room (ER) or hospitalization |

Source: FDA. Guidance for Industry: Toxicity grading scale for healthy adult and adolescent volunteers enrolled in preventive vaccine clinical trials. 2007.

#### Solicited Adverse Events

**Supplementary Table 4.** Solicited Adverse Events by Severity Grades from Days 0 to 7 (Safety Analysis Set) – 3.75 µg.

| Symptoms | Severity | COVLP 3.75 ug<br>unadjuvanted |  | COVLP 3.75 ug<br>with CpG 1018 |  | COVLP 3.75 ug<br>with AS03 |  |
| --- | --- | --- | --- | --- | --- | --- | --- |
|  |  | Vacc. 1 | Vacc 2 | Vacc. 1 | Vacc 2 | Vacc 1 | Vacc 2 |
|  |  | N=20 | N=20 | N=20 | N=20 | N=20 | N=19 |
| Redness at injection site | Grade 1 | 1 (5.0) | 0 (0.0) | 0 (0.0) | 1 (5.0) | 1 (5.0) | 2 (10.5) |
|  | Grade 2 | 0 (0.0) | 0 (0.0) | 0 (0.0) | 0 (0.0) | 0 (0.0) | 3 (15.8) |
|  | Grade 3 | 0 (0.0) | 0 (0.0) | 0 (0.0) | 0 (0.0) | 0 (0.0) | 1 (5.3) |
| Swelling at injection site | Grade 1 | 0 (0.0) | 1 (5.0) | 1 (5.0) | 3 (15.0) | 3 (15.0) | 5 (26.3) |
|  | Grade 2 | 0 (0.0) | 0 (0.0) | 0 (0.0) | 0 (0.0) | 0 (0.0) | 0 (0.0) |
|  | Grade 3 | 0 (0.0) | 0 (0.0) | 0 (0.0) | 0 (0.0) | 0 (0.0) | 1 (5.3) |
| Pain at injection site | Grade 1 | 4 (20.0) | 3 (15.0) | 17 (85.0) | 15 (75.0) | 18 (90.0) | 16 (84.2) |
|  | Grade 2 | 0 (0.0) | 0 (0.0) | 0 (0.0) | 0 (0.0) | 1 (5.0) | 3 (15.8) |
|  | Grade 3 | 0 (0.0) | 0 (0.0) | 0 (0.0) | 0 (0.0) | 0 (0.0) | 0 (0.0) |
| Fever | Grade 1 | 0 (0.0) | 0 (0.0) | 1 (5.0) | 0 (0.0) | 0 (0.0) | 5 (26.3) |
|  | Grade 2 | 0 (0.0) | 0 (0.0) | 0 (0.0) | 0 (0.0) | 0 (0.0) | 2 (10.5) |
|  | Grade 3 | 0 (0.0) | 0 (0.0) | 0 (0.0) | 0 (0.0) | 0 (0.0) | 0 (0.0) |
| Headache | Grade 1 | 3 (15.0) | 2 (10.0) | 9 (45.0) | 6 (30.0) | 6 (30.0) | 6 (31.6) |
|  | Grade 2 | 0 (0.0) | 1 (5.0) | 1 (5.0) | 2 (10.0) | 0 (0.0) | 5 (26.3) |
|  | Grade 3 | 0 (0.0) | 0 (0.0) | 0 (0.0) | 0 (0.0) | 0 (0.0) | 0 (0.0) |
| Muscle aches | Grade 1 | 0 (0.0) | 0 (0.0) | 1 (5.0) | 2 (10.0) | 1 (5.0) | 4 (21.1) |
|  | Grade 2 | 0 (0.0) | 0 (0.0) | 0 (0.0) | 0 (0.0) | 0 (0.0) | 3 (15.8) |
|  | Grade 3 | 0 (0.0) | 0 (0.0) | 0 (0.0) | 0 (0.0) | 0 (0.0) | 0 (0.0) |
| Joint aches | Grade 1 | 0 (0.0) | 0 (0.0) | 0 (0.0) | 0 (0.0) | 1 (5.0) | 1 (5.3) |
|  | Grade 2 | 0 (0.0) | 0 (0.0) | 0 (0.0) | 0 (0.0) | 0 (0.0) | 1 (5.3) |
|  | Grade 3 | 0 (0.0) | 0 (0.0) | 0 (0.0) | 0 (0.0) | 0 (0.0) | 0 (0.0) |
| Fatigue | Grade 1 | 5 (25.0) | 2 (10.0) | 6 (30.0) | 5 (25.0) | 7 (35.0) | 5 (26.3) |
|  | Grade 2 | 0 (0.0) | 0 (0.0) | 1 (5.0) | 0 (0.0) | 0 (0.0) | 5 (26.3) |
|  | Grade 3 | 0 (0.0) | 0 (0.0) | 0 (0.0) | 0 (0.0) | 1 (5.0) | 1 (5.3) |
| Chills | Grade 1 | 1 (5.0) | 1 (5.0) | 1 (5.0) | 0 (0.0) | 0 (0.0) | 8 (42.1) |
|  | Grade 2 | 0 (0.0) | 0 (0.0) | 0 (0.0) | 0 (0.0) | 0 (0.0) | 4 (21.1) |
|  | Grade 3 | 0 (0.0) | 0 (0.0) | 0 (0.0) | 0 (0.0) | 0 (0.0) | 0 (0.0) |
| Discomfort or uneasiness | Grade 1 | 2 (10.0) | 1 (5.0) | 3 (15.0) | 1 (5.0) | 3 (15.0) | 4 (21.1) |
|  | Grade 2 | 0 (0.0) | 0 (0.0) | 0 (0.0) | 0 (0.0) | 0 (0.0) | 5 (26.3) |
|  | Grade 3 | 0 (0.0) | 0 (0.0) | 0 (0.0) | 0 (0.0) | 0 (0.0) | 1 (5.3) |
| Swelling in the neck | Grade 1 | 1 (5.0) | 0 (0.0) | 4 (20.0) | 1 (5.0) | 0 (0.0) | 1 (5.3) |
|  | Grade 2 | 0 (0.0) | 0 (0.0) | 0 (0.0) | 0 (0.0) | 0 (0.0) | 0 (0.0) |
|  | Grade 3 | 0 (0.0) | 0 (0.0) | 0 (0.0) | 0 (0.0) | 0 (0.0) | 0 (0.0) |
| Swelling in the axilla | Grade 1 | 0 (0.0) | 0 (0.0) | 0 (0.0) | 0 (0.0) | 0 (0.0) | 0 (0.0) |
|  | Grade 2 | 0 (0.0) | 0 (0.0) | 0 (0.0) | 0 (0.0) | 0 (0.0) | 0 (0.0) |
|  | Grade 3 | 0 (0.0) | 0 (0.0) | 0 (0.0) | 0 (0.0) | 0 (0.0) | 0 (0.0) |
| Swelling in the groin | Grade 1 | 0 (0.0) | 0 (0.0) | 0 (0.0) | 0 (0.0) | 0 (0.0) | 0 (0.0) |
|  | Grade 2 | 0 (0.0) | 0 (0.0) | 0 (0.0) | 0 (0.0) | 0 (0.0) | 0 (0.0) |
|  | Grade 3 | 0 (0.0) | 0 (0.0) | 0 (0.0) | 0 (0.0) | 0 (0.0) | 0 (0.0) |
| Swelling in the chest wall | Grade 1 | 0 (0.0) | 0 (0.0) | 1 (5.0) | 0 (0.0) | 0 (0.0) | 0 (0.0) |
|  | Grade 2 | 0 (0.0) | 0 (0.0) | 0 (0.0) | 0 (0.0) | 0 (0.0) | 0 (0.0) |
|  | Grade 3 | 0 (0.0) | 0 (0.0) | 0 (0.0) | 0 (0.0) | 0 (0.0) | 0 (0.0) |

**Supplementary Table 5.** Solicited Adverse Events by Severity Grades from Days 0 to 7 (Safety Analysis Set) – 7.5 µg.

| Symptoms | Severity | COVLP 7.5 ug<br>unadjuvanted |  | COVLP 7.5 ug<br>with CpG 1018 |  | COVLP 7.5 ug<br>with AS03 |  |
| --- | --- | --- | --- | --- | --- | --- | --- |
|  |  | Vacc. 1<br>N=20 | Vacc 2<br>N=20 | Vacc. 1<br>N=20 | Vacc 2<br>N=20 | Vacc 1<br>N=20 | Vacc 2<br>N=20 |
| Redness at injection site | Grade 1 | 0 (0.0) | 0 (0.0) | 0 (0.0) | 1 (5.0) | 0 (0.0) | 4 (20.0) |
|  | Grade 2 | 0 (0.0) | 0 (0.0) | 0 (0.0) | 0 (0.0) | 0 (0.0) | 1 (5.0) |
|  | Grade 3 | 0 (0.0) | 0 (0.0) | 0 (0.0) | 0 (0.0) | 0 (0.0) | 2 (10.0) |
| Swelling at injection site | Grade 1 | 0 (0.0) | 1 (5.0) | 4 (20.0) | 3 (15.0) | 6 (30.0) | 5 (25.0) |
|  | Grade 2 | 0 (0.0) | 0 (0.0) | 0 (0.0) | 0 (0.0) | 0 (0.0) | 3 (15.0) |
|  | Grade 3 | 0 (0.0) | 0 (0.0) | 0 (0.0) | 0 (0.0) | 0 (0.0) | 0 (0.0) |
| Pain at injection site | Grade 1 | 6 (30.0) | 2 (10.0) | 15 (75.0) | 12 (60.0) | 15 (75.0) | 14 (70.0) |
|  | Grade 2 | 0 (0.0) | 0 (0.0) | 1 (5.0) | 2 (10.0) | 1 (5.0) | 4 (20.0) |
|  | Grade 3 | 0 (0.0) | 0 (0.0) | 0 (0.0) | 0 (0.0) | 0 (0.0) | 0 (0.0) |
| Fever | Grade 1 | 0 (0.0) | 0 (0.0) | 0 (0.0) | 1 (5.0) | 0 (0.0) | 3 (15.0) |
|  | Grade 2 | 0 (0.0) | 0 (0.0) | 0 (0.0) | 0 (0.0) | 0 (0.0) | 1 (5.0) |
|  | Grade 3 | 0 (0.0) | 0 (0.0) | 0 (0.0) | 0 (0.0) | 0 (0.0) | 0 (0.0) |
| Headache | Grade 1 | 6 (30.0) | 2 (10.0) | 8 (40.0) | 2 (10.0) | 2 (10.0) | 9 (45.0) |
|  | Grade 2 | 0 (0.0) | 2 (10.0) | 0 (0.0) | 1 (5.0) | 0 (0.0) | 6 (30.0) |
|  | Grade 3 | 0 (0.0) | 0 (0.0) | 0 (0.0) | 0 (0.0) | 0 (0.0) | 0 (0.0) |
| Muscle aches | Grade 1 | 1 (5.0) | 1 (5.0) | 1 (5.0) | 2 (10.0) | 0 (0.0) | 8 (40.0) |
|  | Grade 2 | 0 (0.0) | 0 (0.0) | 0 (0.0) | 0 (0.0) | 0 (0.0) | 2 (10.0) |
|  | Grade 3 | 0 (0.0) | 0 (0.0) | 0 (0.0) | 0 (0.0) | 0 (0.0) | 0 (0.0) |
| Joint aches | Grade 1 | 0 (0.0) | 0 (0.0) | 0 (0.0) | 0 (0.0) | 0 (0.0) | 5 (25.0) |
|  | Grade 2 | 0 (0.0) | 1 (5.0) | 0 (0.0) | 0 (0.0) | 0 (0.0) | 2 (10.0) |
|  | Grade 3 | 0 (0.0) | 0 (0.0) | 0 (0.0) | 0 (0.0) | 0 (0.0) | 0 (0.0) |
| Fatigue | Grade 1 | 3 (15.0) | 1 (5.0) | 3 (15.0) | 6 (30.0) | 3 (15.0) | 12 (60.0) |
|  | Grade 2 | 0 (0.0) | 0 (0.0) | 1 (5.0) | 0 (0.0) | 1 (5.0) | 3 (15.0) |
|  | Grade 3 | 0 (0.0) | 0 (0.0) | 0 (0.0) | 1 (5.0) | 0 (0.0) | 0 (0.0) |
| Chills | Grade 1 | 1 (5.0) | 1 (5.0) | 1 (5.0) | 4 (20.0) | 0 (0.0) | 9 (45.0) |
|  | Grade 2 | 0 (0.0) | 0 (0.0) | 0 (0.0) | 0 (0.0) | 0 (0.0) | 3 (15.0) |
|  | Grade 3 | 0 (0.0) | 0 (0.0) | 0 (0.0) | 0 (0.0) | 0 (0.0) | 0 (0.0) |
| Discomfort or uneasiness | Grade 1 | 0 (0.0) | 0 (0.0) | 2 (10.0) | 2 (10.0) | 0 (0.0) | 8 (40.0) |
|  | Grade 2 | 0 (0.0) | 0 (0.0) | 0 (0.0) | 0 (0.0) | 0 (0.0) | 3 (15.0) |
|  | Grade 3 | 0 (0.0) | 0 (0.0) | 0 (0.0) | 0 (0.0) | 0 (0.0) | 0 (0.0) |
| Swelling in the neck | Grade 1 | 1 (5.0) | 0 (0.0) | 0 (0.0) | 1 (5.0) | 1 (5.0) | 2 (10.0) |
|  | Grade 2 | 0 (0.0) | 0 (0.0) | 0 (0.0) | 0 (0.0) | 0 (0.0) | 0 (0.0) |
|  | Grade 3 | 0 (0.0) | 0 (0.0) | 0 (0.0) | 0 (0.0) | 0 (0.0) | 0 (0.0) |
| Swelling in the axilla | Grade 1 | 1 (5.0) | 0 (0.0) | 0 (0.0) | 0 (0.0) | 0 (0.0) | 1 (5.0) |
|  | Grade 2 | 0 (0.0) | 0 (0.0) | 0 (0.0) | 0 (0.0) | 0 (0.0) | 0 (0.0) |
|  | Grade 3 | 0 (0.0) | 0 (0.0) | 0 (0.0) | 0 (0.0) | 0 (0.0) | 0 (0.0) |
| Swelling in the groin | Grade 1 | 1 (5.0) | 0 (0.0) | 0 (0.0) | 0 (0.0) | 0 (0.0) | 1 (5.0) |
|  | Grade 2 | 0 (0.0) | 0 (0.0) | 0 (0.0) | 0 (0.0) | 0 (0.0) | 0 (0.0) |
|  | Grade 3 | 0 (0.0) | 0 (0.0) | 0 (0.0) | 0 (0.0) | 0 (0.0) | 0 (0.0) |
| Swelling in the chest wall | Grade 1 | 0 (0.0) | 0 (0.0) | 0 (0.0) | 0 (0.0) | 0 (0.0) | 1 (5.0) |
|  | Grade 2 | 0 (0.0) | 0 (0.0) | 0 (0.0) | 0 (0.0) | 0 (0.0) | 0 (0.0) |
|  | Grade 3 | 0 (0.0) | 0 (0.0) | 0 (0.0) | 0 (0.0) | 0 (0.0) | 0 (0.0) |

**Supplementary Table 6.** Solicited Adverse Events by Severity Grades from Days 0 to 7 (Safety Analysis Set) – 15 µg.

| Symptoms | Severity | COVLP 15 ug<br>unadjuvanted |  | COVLP 15 ug<br>with CpG 1018 |  | COVLP 15 ug<br>with AS03 |  |
| --- | --- | --- | --- | --- | --- | --- | --- |
|  |  | Vacc 1<br>N=20 | Vacc 2<br>N=20 | Vacc. 1<br>N=20 | Vacc 2<br>N=19 | Vacc 1<br>N=20 | Vacc 2<br>N=20 |
| Redness at injection site | Grade 1 | 0 (0.0) | 0 (0.0) | 0 (0.0) | 0 (0.0) | 1 (5.0) | 3 (15.0) |
|  | Grade 2 | 0 (0.0) | 0 (0.0) | 0 (0.0) | 0 (0.0) | 0 (0.0) | 1 (5.0) |
|  | Grade 3 | 0 (0.0) | 0 (0.0) | 0 (0.0) | 0 (0.0) | 0 (0.0) | 0 (0.0) |
| Swelling at injection site | Grade 1 | 0 (0.0) | 0 (0.0) | 2 (10.0) | 1 (5.3) | 3 (15.0) | 3 (15.0) |
|  | Grade 2 | 0 (0.0) | 0 (0.0) | 0 (0.0) | 0 (0.0) | 1 (5.0) | 1 (5.0) |
|  | Grade 3 | 0 (0.0) | 0 (0.0) | 0 (0.0) | 0 (0.0) | 0 (0.0) | 1 (5.0) |
| Pain at injection site | Grade 1 | 6 (30.0) | 7 (35.0) | 16 (80.0) | 13 (68.4) | 13 (65.0) | 14 (70.0) |
|  | Grade 2 | 0 (0.0) | 1 (5.0) | 0 (0.0) | 1 (5.3) | 4 (20.0) | 2 (10.0) |
|  | Grade 3 | 0 (0.0) | 0 (0.0) | 0 (0.0) | 0 (0.0) | 0 (0.0) | 0 (0.0) |
| Fever | Grade 1 | 0 (0.0) | 0 (0.0) | 0 (0.0) | 1 (5.3) | 0 (0.0) | 5 (25.0) |
|  | Grade 2 | 0 (0.0) | 0 (0.0) | 0 (0.0) | 0 (0.0) | 0 (0.0) | 1 (5.0) |
|  | Grade 3 | 0 (0.0) | 0 (0.0) | 0 (0.0) | 0 (0.0) | 0 (0.0) | 0 (0.0) |
| Headache | Grade 1 | 3 (15.0) | 2 (10.0) | 6 (30.0) | 4 (21.1) | 7 (35.0) | 6 (30.0) |
|  | Grade 2 | 1 (5.0) | 0 (0.0) | 0 (0.0) | 0 (0.0) | 1 (5.0) | 3 (15.0) |
|  | Grade 3 | 0 (0.0) | 0 (0.0) | 0 (0.0) | 0 (0.0) | 0 (0.0) | 0 (0.0) |
| Muscle aches | Grade 1 | 1 (5.0) | 0 (0.0) | 2 (10.0) | 1 (5.3) | 1 (5.0) | 2 (10.0) |
|  | Grade 2 | 0 (0.0) | 0 (0.0) | 0 (0.0) | 0 (0.0) | 0 (0.0) | 3 (15.0) |
|  | Grade 3 | 0 (0.0) | 0 (0.0) | 0 (0.0) | 0 (0.0) | 0 (0.0) | 0 (0.0) |
| Joint aches | Grade 1 | 0 (0.0) | 0 (0.0) | 1 (5.0) | 0 (0.0) | 1 (5.0) | 2 (10.0) |
|  | Grade 2 | 0 (0.0) | 0 (0.0) | 0 (0.0) | 0 (0.0) | 0 (0.0) | 1 (5.0) |
|  | Grade 3 | 0 (0.0) | 0 (0.0) | 0 (0.0) | 0 (0.0) | 0 (0.0) | 0 (0.0) |
| Fatigue | Grade 1 | 0 (0.0) | 1 (5.0) | 0 (0.0) | 6 (31.6) | 4 (20.0) | 9 (45.0) |
|  | Grade 2 | 1 (5.0) | 0 (0.0) | 1 (5.0) | 0 (0.0) | 0 (0.0) | 2 (10.0) |
|  | Grade 3 | 0 (0.0) | 0 (0.0) | 0 (0.0) | 0 (0.0) | 0 (0.0) | 0 (0.0) |
| Chills | Grade 1 | 0 (0.0) | 0 (0.0) | 0 (0.0) | 2 (10.5) | 1 (5.0) | 6 (30.0) |
|  | Grade 2 | 0 (0.0) | 0 (0.0) | 0 (0.0) | 0 (0.0) | 0 (0.0) | (10.0) |
|  | Grade 3 | 0 (0.0) | 0 (0.0) | 0 (0.0) | 0 (0.0) | 0 (0.0) | 0 (0.0) |
| Discomfort or uneasiness | Grade 1 | 0 (0.0) | 0 (0.0) | 1 (5.0) | 2 (10.5) | 2 (10.0) | 4 (20.0) |
|  | Grade 2 | 0 (0.0) | 0 (0.0) | 1 (5.0) | 0 (0.0) | 0 (0.0) | 2 (10.0) |
|  | Grade 3 | 0 (0.0) | 0 (0.0) | 0 (0.0) | 0 (0.0) | 0 (0.0) | 0 (0.0) |
| Swelling in the neck | Grade 1 | 2 (10.0) | 0 (0.0) | 1 (5.0) | 1 (5.3) | 2 (10.0) | 3 (15.0) |
|  | Grade 2 | 0 (0.0) | 0 (0.0) | 0 (0.0) | 0 (0.0) | 0 (0.0) | 0 (0.0) |
|  | Grade 3 | 0 (0.0) | 0 (0.0) | 0 (0.0) | 0 (0.0) | 0 (0.0) | 0 (0.0) |
| Swelling in the axilla | Grade 1 | 0 (0.0) | 0 (0.0) | 0 (0.0) | 1 (5.3) | 2 (10.0) | 2 (10.0) |
|  | Grade 2 | 0 (0.0) | 0 (0.0) | 0 (0.0) | 0 (0.0) | 0 (0.0) | 0 (0.0) |
|  | Grade 3 | 0 (0.0) | 0 (0.0) | 0 (0.0) | 0 (0.0) | 0 (0.0) | 0 (0.0) |
| Swelling in the groin | Grade 1 | 0 (0.0) | 0 (0.0) | 0 (0.0) | 0 (0.0) | 0 (0.0) | 0 (0.0) |
|  | Grade 2 | 0 (0.0) | 0 (0.0) | 0 (0.0) | 0 (0.0) | 0 (0.0) | 0 (0.0) |
|  | Grade 3 | 0 (0.0) | 0 (0.0) | 0 (0.0) | 0 (0.0) | 0 (0.0) | 0 (0.0) |
| Swelling in the chest wall | Grade 1 | 0 (0.0) | 0 (0.0) | 1 (5.0) | 0 (0.0) | 0 (0.0) | 0 (0.0) |
|  | Grade 2 | 0 (0.0) | 0 (0.0) | 0 (0.0) | 0 (0.0) | 0 (0.0) | 0 (0.0) |
|  | Grade 3 | 0 (0.0) | 0 (0.0) | 0 (0.0) | 0 (0.0) | 0 (0.0) | 0 (0.0) |

#### Unsolicited Adverse Events

**Supplementary Table 7.** Treatment-Emergent Unsolicited Adverse Events up to 21 days after last vaccination (Safety Analysis Set) – Dose 3.75 µg.

| **Adverse Event**<br>**SOC/PT** | **COVLP 3.75 ug**<br>**unadjuvanted**<br>**(N=20)** | **COVLP 3.75 ug**<br>**with CpG 1018**<br>**(N=20)** | **COVLP 3.75 ug**<br>**with AS03**<br>**(N=20)** |
| --- | --- | --- | --- |
| Subjects with at least one unsolicited AE | 8 | 9 | 8 |
| Eye disorders |  |  |  |
| Conjunctivitis allergic | 1 (5.0) | 0 (0.0) | 0 (0.0) |
| Gastrointestinal disorders |  |  |  |
| Abdominal pain upper | 0 (0.0) | 1 (5.0) | 0 (0.0) |
| General disorders and administration site conditions |  |  |  |
| Chest discomfort | 1 (5.0) | 0 (0.0) | 0 (0.0) |
| Fatigue | 0 (0.0) | 1 (5.0) | 0 (0.0) |
| Feeling hot | 0 (0.0) | 0 (0.0) | 1 (5.0) |
| Injection site pain | 0 (0.0) | 0 (0.0) | 1 (5.0) |
| Injection site swelling | 0 (0.0) | 0 (0.0) | 1 (5.0) |
| Infections and infestations |  |  |  |
| Bacterial vaginosis | 0 (0.0) | 0 (0.0) | 1 (5.0) |
| Hordeolum | 0 (0.0) | 1 (5.0) | 0 (0.0) |
| Vaginal infection | 0 (0.0) | 0 (0.0) | 1 (5.0) |
| Injury, poisoning and procedural complications |  |  |  |
| Ligament sprain | 1 (5.0) | 0 (0.0) | 0 (0.0) |
| Tooth fracture | 1 (5.0) | 0 (0.0) | 0 (0.0) |
| Investigations |  |  |  |
| Blood glucose increased | 0 (0.0) | 1 (5.0) | 0 (0.0) |
| Musculoskeletal and connective tissue disorders |  |  |  |
| Plantar fasciitis | 0 (0.0) | 1 (5.0) | 0 (0.0) |
| Nervous system disorders |  |  |  |
| Dizziness exertional | 0 (0.0) | 1 (5.0) | 0 (0.0) |
| Paraesthesia | 1 (5.0) | 0 (0.0) | 0 (0.0) |
| Psychiatric disorders |  |  |  |
| Depressed mood | 0 (0.0) | 1 (5.0) | 0 (0.0) |
| Irritability | 0 (0.0) | 1 (5.0) | 0 (0.0) |
| Reproductive system and breast disorders |  |  |  |
| Dysmenorrhoea | 0 (0.0) | 0 (0.0) | 1 (5.0) |
| Respiratory, thoracic and mediastinal disorders |  |  |  |
| Asthma exercise induced | 0 (0.0) | 0 (0.0) | 1 (5.0) |
| Skin and subcutaneous tissue disorders |  |  |  |
| Rash maculo-papular | 1 (5.0) | 0 (0.0) | 0 (0.0) |

**Supplementary Table 8.** Treatment-Emergent Unsolicited Adverse Events up to 21 days after last vaccination (Safety Analysis Set) – Dose 7.5 µg.

| **Adverse Event**<br>**SOC/PT** | **COVLP 7.5 ug**<br>**unadjuvanted**<br>**(N=20)** | **COVLP 7.5 ug**<br>**with CpG 1018**<br>**(N=20)** | **COVLP 7.5 ug**<br>**with AS03**<br>**(N=20)** |
| --- | --- | --- | --- |
| Subjects with at least one unsolicited AE | 9 | 11 | 7 |
| Blood and lymphatic system disorders |  |  |  |
| Lymphadenopathy | 0 (0.0) | 0 (0.0) | 1 (5.0) |
| Gastrointestinal disorders |  |  |  |
| Diarrhoea | 0 (0.0) | 0 (0.0) | 1 (5.0) |
| General disorders and administration site conditions |  |  |  |
| Injection site papule | 0 (0.0) | 0 (0.0) | 1 (5.0) |
| Investigations |  |  |  |
| Blood pressure decreased | 1 (5.0) | 0 (0.0) | 0 (0.0) |
| Blood pressure increased | 1 (5.0) | 0 (0.0) | 0 (0.0) |
| Metabolism and nutrition disorders |  |  |  |
| Hypercholesterolaemia | 1 (5.0) | 0 (0.0) | 0 (0.0) |
| Musculoskeletal and connective tissue disorders |  |  |  |
| Costochondritis | 0 (0.0) | 1 (5.0) | 0 (0.0) |
| Limb discomfort | 0 (0.0) | 1 (5.0) | 0 (0.0) |
| Pain in extremity | 0 (0.0) | 0 (0.0) | 1 (5.0) |
| Nervous system disorders |  |  |  |
| Dizziness | 0 (0.0) | 1 (5.0) | 0 (0.0) |
| Psychiatric disorders |  |  |  |
| Anxiety | 0 (0.0) | 1 (5.0) | 0 (0.0) |
| Reproductive system and breast disorders |  |  |  |
| Vulvovaginal pruritus | 0 (0.0) | 1 (5.0) | 0 (0.0) |
| Respiratory, thoracic and mediastinal disorders |  |  |  |
| Dry throat | 1 (5.0) | 0 (0.0) | 0 (0.0) |
| Rhinitis allergic | 1 (5.0) | 0 (0.0) | 0 (0.0) |
| Throat irritation | 0 (0.0) | 2 (10.0) | 0 (0.0) |
| Skin and subcutaneous tissue disorders |  |  |  |
| Dermatitis contact | 0 (0.0) | 1 (5.0) | 0 (0.0) |
| Erythema | 0 (0.0) | 1 (5.0) | 0 (0.0) |
| Rash | 1 (5.0) | 0 (0.0) | 0 (0.0) |

**Supplementary Table 9.** Treatment-Emergent Unsolicited Adverse Events up to 21 days after last vaccination (Safety Analysis Set) – Dose 15 µg.

| **Adverse Event**<br>**SOC/PT** | **COVLP 15 ug**<br>**unadjuvanted**<br>**(N=20)** | **COVLP 15 ug**<br>**with CpG 1018**<br>**(N=20)** | **COVLP 15 ug**<br>**with AS03**<br>**(N=20)** |
| --- | --- | --- | --- |
| Subjects with at least one unsolicited AE | 8 | 6 | 11 |
| Ear and labyrinth disorders |  |  |  |
| Ear pain | 0 (0.0) | 0 (0.0) | 1 (5.0) |
| Gastrointestinal disorders |  |  |  |
| Abdominal discomfort | 0 (0.0) | 1 (5.0) | 0 (0.0) |
| Abdominal pain | 1 (5.0) | 0 (0.0) | 0 (0.0) |
| Nausea | 0 (0.0) | 0 (0.0) | 1 (5.0) |
| Toothache | 1 (5.0) | 0 (0.0) | 0 (0.0) |
| Vomiting | 0 (0.0) | 0 (0.0) | 2 (10.0) |
| General disorders and administration site conditions |  |  |  |
| Feeling cold | 0 (0.0) | 0 (0.0) | 1 (5.0) |
| Injection site bruising | 0 (0.0) | 0 (0.0) | 1 (5.0) |
| Injection site erythema | 0 (0.0) | 0 (0.0) | 1 (5.0) |
| Injection site warmth | 0 (0.0) | 0 (0.0) | 1 (5.0) |
| Swelling | 0 (0.0) | 0 (0.0) | 1 (5.0) |
| Infections and infestations |  |  |  |
| Oral herpes | 1 (5.0) | 0 (0.0) | 0 (0.0) |
| Injury, poisoning and procedural complications |  |  |  |
| Joint injury | 0 (0.0) | 1 (5.0) | 0 (0.0) |
| Scratch | 0 (0.0) | 1 (5.0) | 0 (0.0) |
| Wound | 0 (0.0) | 1 (5.0) | 0 (0.0) |
| Investigations |  |  |  |
| Aspartate aminotransferase increased | 0 (0.0) | 0 (0.0) | 1 (5.0) |
| Blood cholesterol increased | 1 (5.0) | 0 (0.0) | 0 (0.0) |
| Blood creatine phosphokinase increased | 0 (0.0) | 0 (0.0) | 2 (10.0) |
| Musculoskeletal and connective tissue disorders |  |  |  |
| Back pain | 0 (0.0) | 0 (0.0) | 1 (5.0) |
| Muscle spasms | 0 (0.0) | 0 (0.0) | 1 (5.0) |
| Nervous system disorders |  |  |  |
| Headache | 0 (0.0) | 0 (0.0) | 1 (5.0) |
| Presyncope | 2 (10.0) | 0 (0.0) | 0 (0.0) |
| Taste disorder | 0 (0.0) | 0 (0.0) | 1 (5.0) |
| Psychiatric disorders |  |  |  |
| Insomnia | 0 (0.0) | 0 (0.0) | 1 (5.0) |
| Respiratory, thoracic and mediastinal disorders |  |  |  |
| Oropharyngeal pain | 0 (0.0) | 0 (0.0) | 2 (10.0) |
| Rhinorrhoea | 0 (0.0) | 1 (5.0) | 0 (0.0) |
| Skin and subcutaneous tissue disorders |  |  |  |
| Dyshidrotic eczema | 1 (5.0) | 0 (0.0) | 0 (0.0) |
| Vascular disorders |  |  |  |
| Hot flush | 0 (0.0) | 1 (5.0) | 0 (0.0) |

#### Trial Profile

**Supplementary Figure 1.**
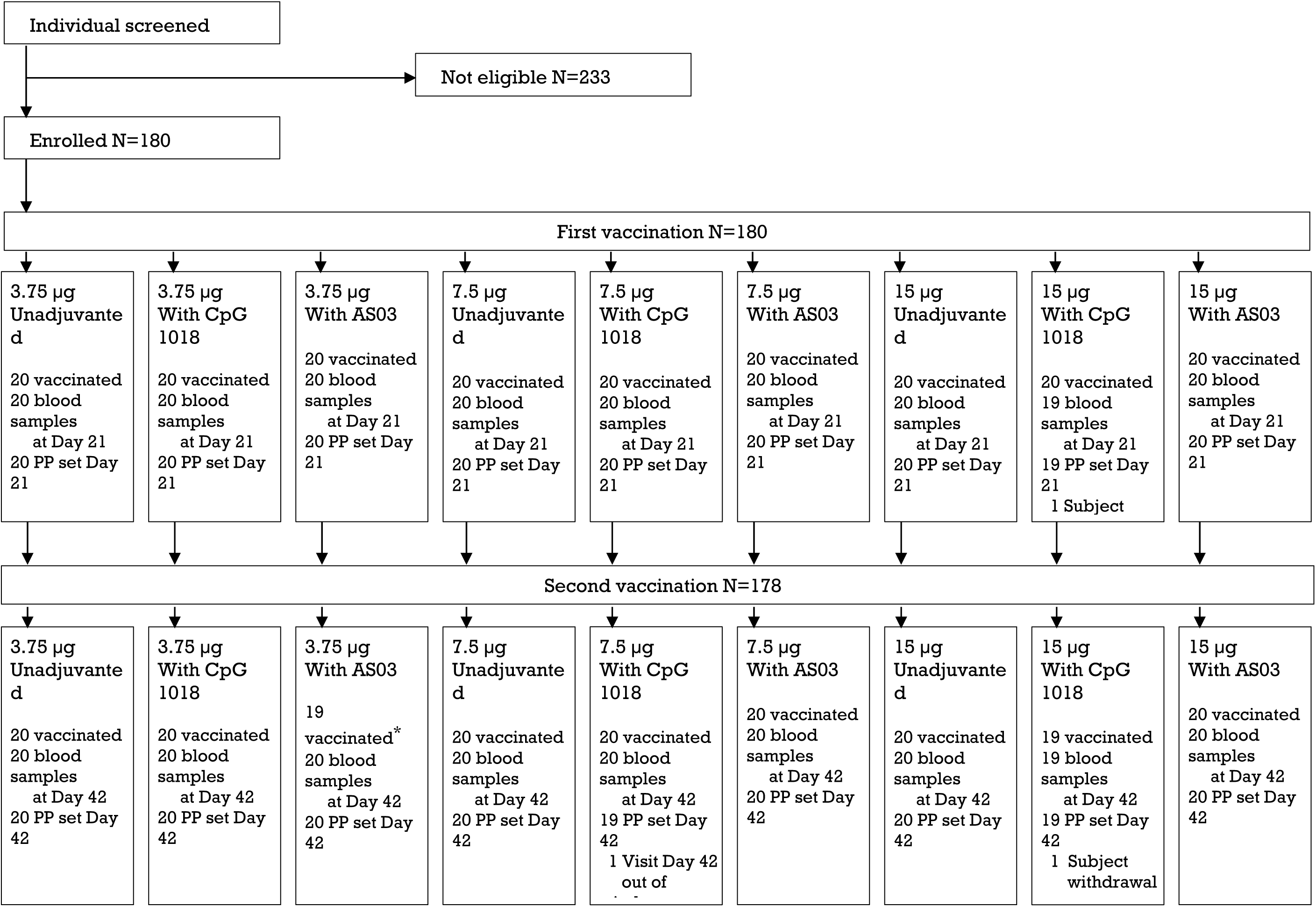
Consort Flow Diagram. *One subject did not receive the second vaccination following Grade 3 adverse event but accepted to have blood collection for immunogenicity.

#### Detailed Immunogenicity Results

**Supplementary Table 10.**
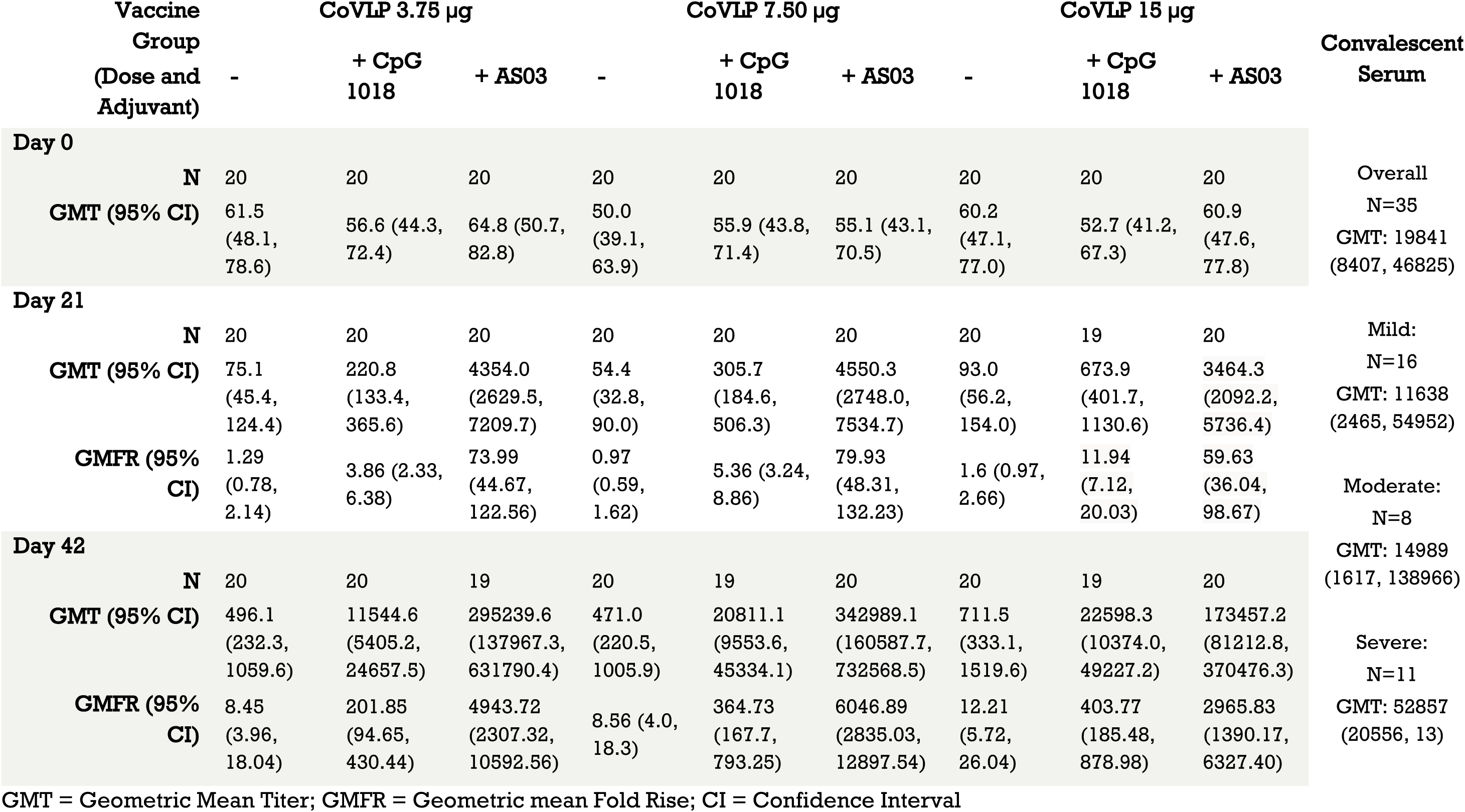
CoVLP-induced Binding Antibody IgG Titers (Anti-Spike ELISA)

| Vaccine Group<br>(Dose and Adjuvant) | CoVLP 3.75 µg |  |  | CoVLP 7.50 µg |  |  | CoVLP 15 µg |  |  | Convalescent Serum |
| --- | --- | --- | --- | --- | --- | --- | --- | --- | --- | --- |
|  | - | + CpG 1018 | + AS03 | - | + CpG 1018 | + AS03 | - | + CpG 1018 | + AS03 |  |
| Day 0 |  |  |  |  |  |  |  |  |  |  |
| N | 20 | 20 | 20 | 20 | 20 | 20 | 20 | 20 | 20 | Overall |
| GMT (95% CI) | 61.5<br>(48.1, 78.6) | 56.6 (44.3, 72.4) | 64.8 (50.7, 82.8) | 50.0<br>(39.1, 63.9) | 55.9 (43.8, 71.4) | 55.1 (43.1, 70.5) | 60.2<br>(47.1, 77.0) | 52.7 (41.2, 67.3) | 60.9<br>(47.6, 77.8) | N=35<br>GMT: 19841<br>(8407, 46825) |
| Day 21 |  |  |  |  |  |  |  |  |  |  |
| N | 20 | 20 | 20 | 20 | 20 | 20 | 20 | 19 | 20 | Mild: |
| GMT (95% CI) | 75.1<br>(45.4, 124.4) | 220.8<br>(133.4, 365.6) | 4354.0<br>(2629.5, 7209.7) | 54.4<br>(32.8, 90.0) | 305.7<br>(184.6, 506.3) | 4550.3<br>(2748.0, 7534.7) | 93.0<br>(56.2, 154.0) | 673.9<br>(401.7, 1130.6) | 3464.3<br>(2092.2, 5736.4) | N=16<br>GMT: 11638<br>(2465, 54952) |
| GMFR (95% CI) | 1.29<br>(0.78, 2.14) | 3.86 (2.33, 6.38) | 73.99<br>(44.67, 122.56) | 0.97<br>(0.59, 1.62) | 5.36 (3.24, 8.86) | 79.93<br>(48.31, 132.23) | 1.6 (0.97, 2.66) | 11.94<br>(7.12, 20.03) | 59.63<br>(36.04, 98.67) | Moderate:<br>N=8<br>GMT: 14989<br>(1617, 138966) |
| Day 42 |  |  |  |  |  |  |  |  |  |  |
| N | 20 | 20 | 19 | 20 | 19 | 20 | 20 | 19 | 20 | Severe: |
| GMT (95% CI) | 496.1<br>(232.3, 1059.6) | 11544.6<br>(5405.2, 24657.5) | 295239.6<br>(137967.3, 631790.4) | 471.0<br>(220.5, 1005.9) | 20811.1<br>(9553.6, 45334.1) | 342989.1<br>(160587.7, 732568.5) | 711.5<br>(333.1, 1519.6) | 22598.3<br>(10374.0, 49227.2) | 173457.2<br>(81212.8, 370476.3) | N=11<br>GMT: 52857<br>(20556, 13) |
| GMFR (95% CI) | 8.45<br>(3.96, 18.04) | 201.85<br>(94.65, 430.44) | 4943.72<br>(2307.32, 10592.56) | 8.56 (4.0, 18.3) | 364.73<br>(167.7, 793.25) | 6046.89<br>(2835.03, 12897.54) | 12.21<br>(5.72, 26.04) | 403.77<br>(185.48, 878.98) | 2965.83<br>(1390.17, 6327.40) |  |

GMT = Geometric Mean Titer; GMFR = Geometric mean Fold Rise; CI = Confidence Interval

**Supplementary Table 11.** CoVLP-induced Neutralization Antibody (Nab) Titers (Pseudovirion Neutralization Assay)

| Vaccine Group<br>(Dose and Adjuvant) | CoVLP 3.75 µg |  |  | CoVLP 7.50 µg |  |  | CoVLP 15 µg |  |  | Convalescent Serum |
| --- | --- | --- | --- | --- | --- | --- | --- | --- | --- | --- |
|  | - | + CpG 1018 | + AS03 | - | + CpG 1018 | + AS03 | - | + CpG 1018 | + AS03 |  |
| Day 0 |  |  |  |  |  |  |  |  |  |  |
| N | 20 | 20 | 20 | 20 | 20 | 20 | 20 | 20 | 20 |  |
| GMT (95% CI) | 5.0 (5.0, 5.0) | 5.0 (5.0, 5.0) | 5.0 (5.0, 5.0) | 5.0 (5.0, 5.0) | 5.0 (5.0, 5.0) | 5.0 (5.0, 5.0) | 5.0 (5.0, 5.0) | 5.0 (5.0, 5.0) | 5.0 (5.0, 5.0) |  |
| Day 21 |  |  |  |  |  |  |  |  |  |  |
| N | 20 | 20 | 20 | 20 | 19 | 19 | 19 | 18 | 20 | Overall<br>N=24<br>GMT: 86.14 (42.0, 176.7)<br>Mild: N=16<br>GMT: 91.0 (34.5, 239.7) |
| GMT (95% CI) | 5.0 (3.2-7.7) | 5.4 (3.5-8.3) | 41.6 (27.0, 64.1) | 5.0 (3.2, 7.7) | 5.7 (3.6, 8.8) | 37.8 (24.3, 58.9) | 5.0 (3.2, 7.8) | 5.5 (3.5, 8.7) | 23.6 (15.3, 36.4) |  |
| GMFR (95% CI) | 1.00 (0.65, 1.54) | 1.07 (0.70, 1.65) | 8.33 (5.41, 12.82) | 1.0 (0.65, 1.54) | 1.13 (0.73, 1.77) | 7.57 (4.86, 11.78) | 1.0 (0.64, 1.56) | 1.1 (0.7, 1.73) | 4.72 (3.07, 7.27) |  |
| SCR (%) (95%CI) | 0 (0.0%) (0.0, 16.8) | 0 (0.0%) (0.0, 16.8) | 12 (60.0%) (36.1, 80.9) | 0 (0.0%) (0.0, 16.8) | 1 (5.3%) (0.1, 26.0) | 8 (42.1%) (20.3, 66.5) | 0 (0.0%) (0.0, 17.6) | 0 (0.0%) (0.0, 18.5) | 9 (45.0%) (23.1, 68.5) |  |
| Day 42 |  |  |  |  |  |  |  |  |  |  |
| N | 20 | 19 | 18 | 20 | 19 | 19 | 20 | 16 | 20 | Moderate: N=8<br>GMT: 77.2 (21.1, 283.1) |
| GMT (95% CI) | 7.9 (4.5-13.7) | 71.3 (40.4, 125.9) | 2118.3 (1228.7, 3651.9) | 9.8 (5.7, 17.1) | 112.2 (63.6, 198.0) | 1883.8 (1082.9, 3277.0) | 11.2 (6.4, 19.4) | 118.1 (63.6, 219.4) | 1200.9 (690.3, 2089.0) |  |
| GMFR (95% CI) | 1.57 (0.90, 2.74) | 14.26 (8.08, 25.17) | 423.66 (245.75, 730.39) | 1.97 (1.13, 3.42) | 22.44 (12.71, 39.60) | 376.76 (216.58, 655.39) | 2.23 (1.28, 3.88) | 23.62 (12.72, 43.87) | 240.18 (138.07, 417.81) |  |
| SCR (%) (95%CI) | 2 (10.0%) (1.2, 31.7) | 12 (63.2%) (38.4, 83.7) | 18 (100%) (81.5, 100) | 4 (20.0%) (5.7, 43.7) | 14 (73.7%) (48.8, 90.9) | 20 (100.0%) (83.2, 100.0) | 5 (25.0%) (8.7, 49.1) | 12 (75.0%) (47.6, 92.7) | 20 (100.0%) (83.2, 100.0) |  |

GMT = Geometric Mean Titer; GMFR = Geometric mean Fold Rise; SCR = Seroconversion ratio; CI = Confidence Interval

**Supplementary Table 12.**
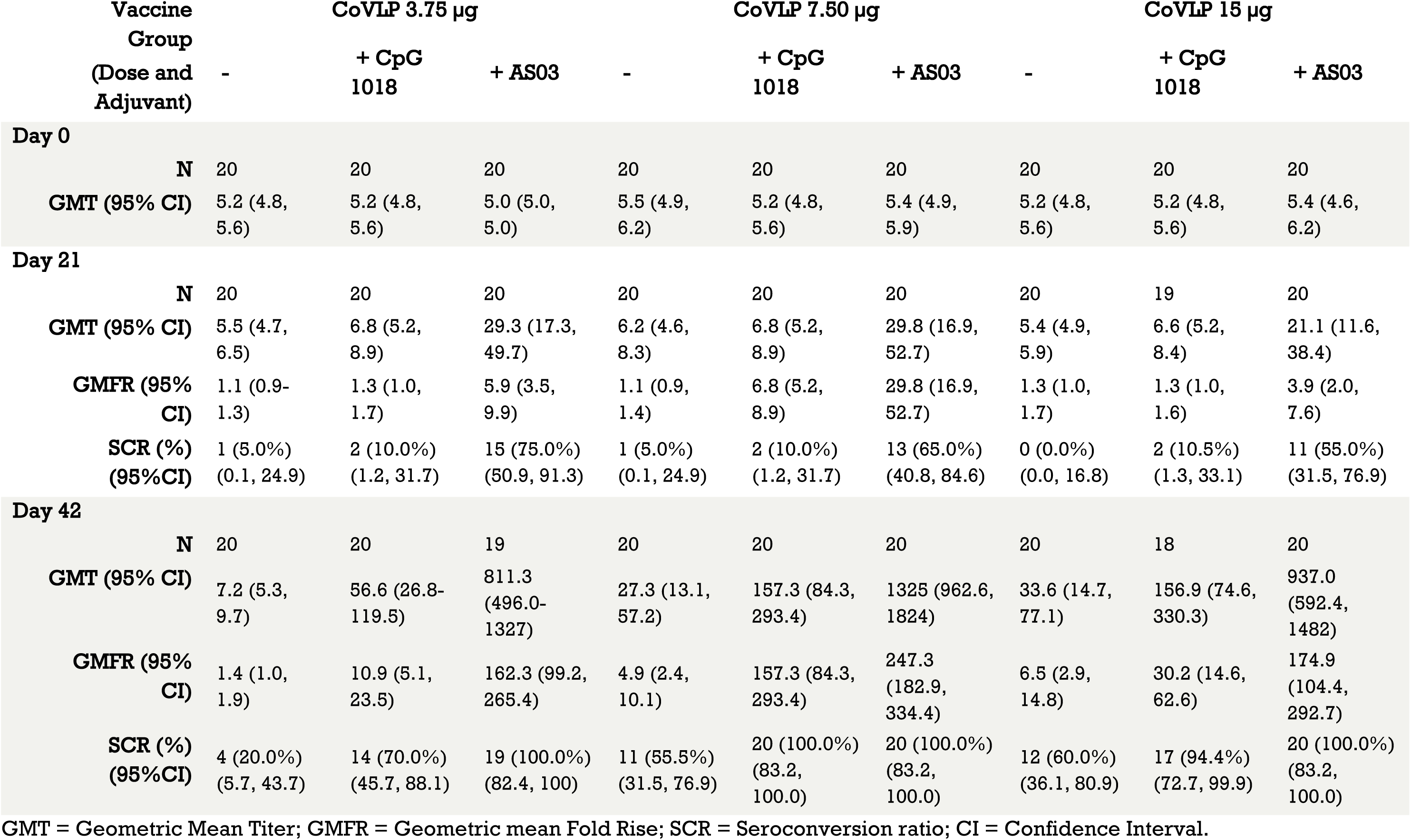
CoVLP-induced Neutralization Antibody (Nab) Titers (Live Wild-Type Virus Neutralization Assay)

**Supplementary Table 13.**
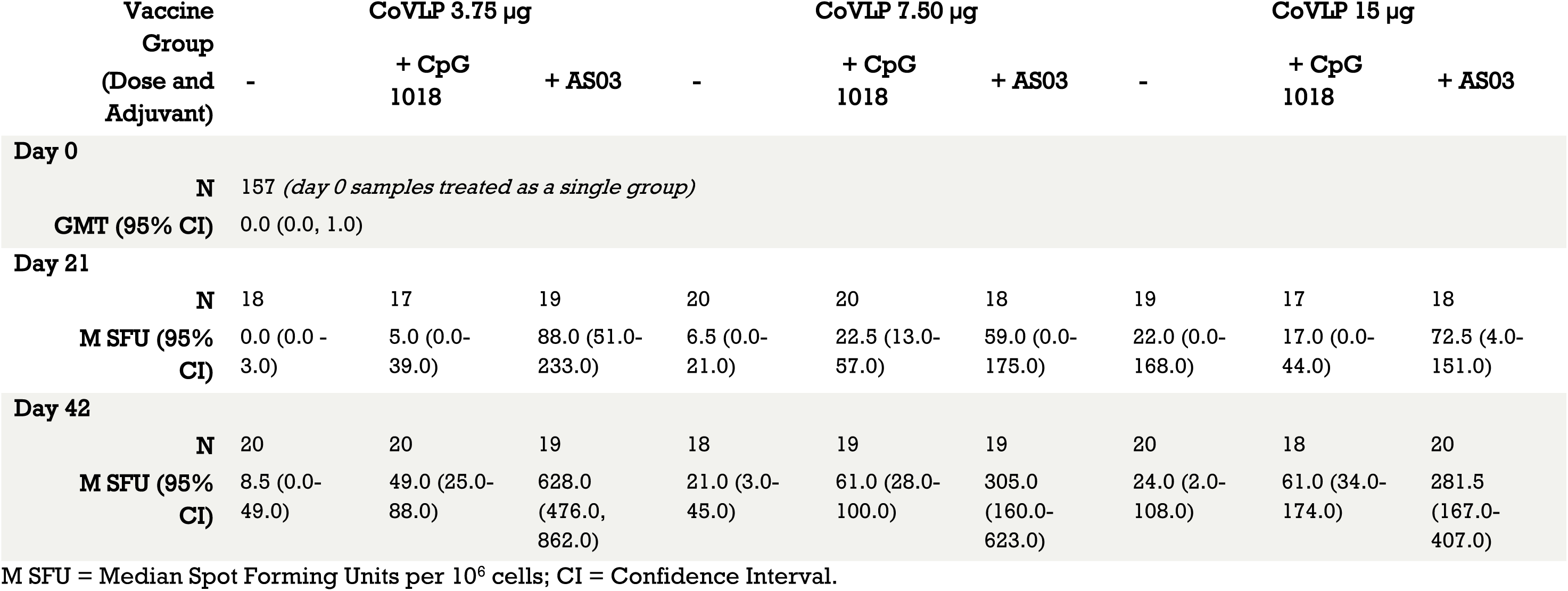
CoVLP-induced IFNγ Cellular Response (ELISpot)

**Supplementary Table 14.**
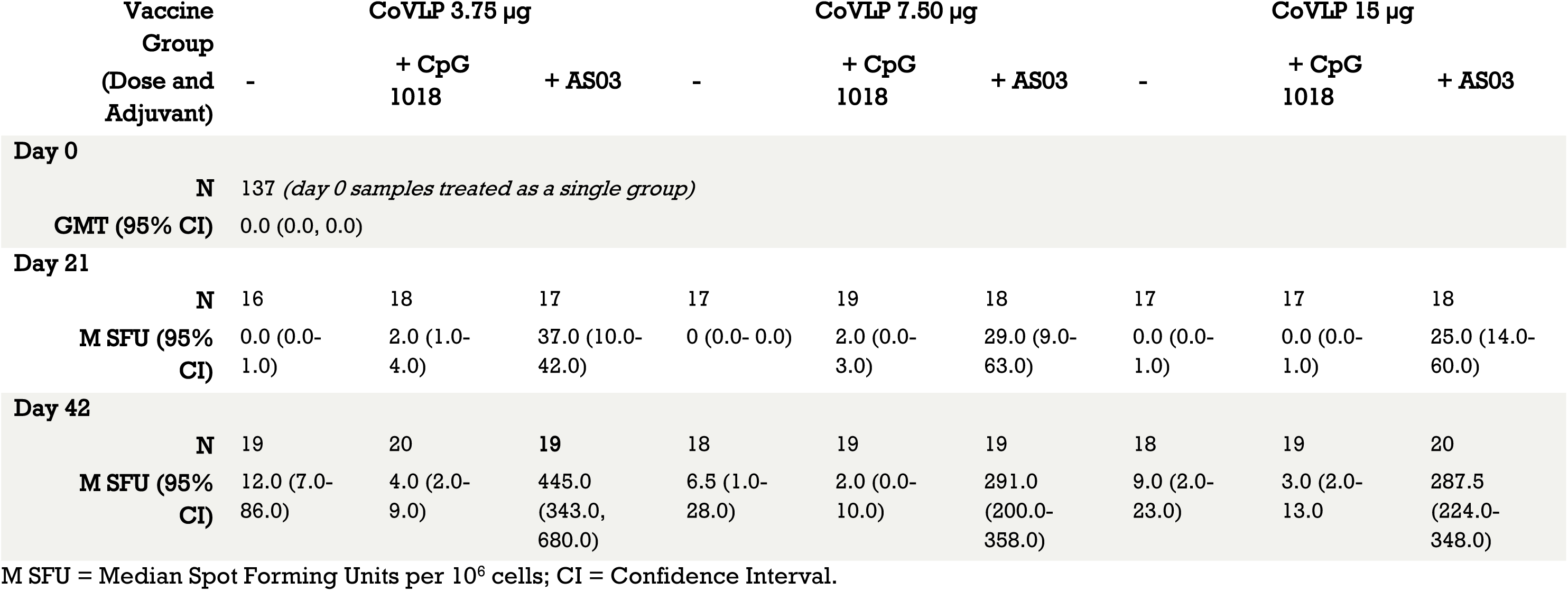
CoVLP-induced IL4 Cellular Response (ELISpot)

## References

1. Guan WJ, Ni ZY, Hu Y, et al. Clinical Characteristics of Coronavirus Disease 2019 in China. Engl J Med 2020;382:1708–20.

2. Zhu F-C, Guan X-H, Li Y-H, et al. Immunogenicity and safety of a recombinant adenovirus type-5-vectored COVID-19 vaccine in healthy adults aged 18 years or older: a randomised, double-blind, placebo-controlled, phase 2 trial. The Lancet 2020;396:479–88.

3. World Health Organization, WHO Director-General’s opening remarks at the media briefing on COVID-19 - 11 March 2020. 2020. at https://www.who.int/dg/speeches/detail/who-director-general-s-opening-remarks-at-the-media-briefing-on-covid-1911-march-2020.)

4. World Health Organization, Coronavirus disease (COVID-19) Weekly Epidemiological Update. 2020. at https://www.who.int/docs/default-source/coronaviruse/situation-reports/20201012-weekly-epi-update-9.pdf.)

5. Santos J, Brierley S, Gandhi MJ, Cohen MA, Moschella PC, Declan ABL. Repurposing Therapeutics for Potential Treatment of SARS-CoV-2: A Review. Viruses 2020;12.

6. Lu H. Drug treatment options for the 2019-new coronavirus (2019-nCoV). Biosci Trends 2020;14:69–71.

7. Wiersinga WJ, Rhodes A, Cheng AC, Peacock SJ, Prescott HC. Pathophysiology, Transmission, Diagnosis, and Treatment of Coronavirus Disease 2019 (COVID-19): A Review. Jama 2020;324:782–93.

8. de Wit E, van Doremalen N, Falzarano D, Munster VJ. SARS and MERS: recent insights into emerging coronaviruses. Nat Rev Microbiol 2016;14:523–34.

9. Burki T. CEPI: preparing for the worst. Lancet Infect Dis 2017;17:265–6.

10. World Health Organization, Draft landscape of COVID-19 candidate vaccines. 2020. at https://www.who.int/publications/m/item/draft-landscape-of-covid-19-candidate-vaccines.)

11. Thanh Le T, Andreadakis Z, Kumar A, et al. The COVID-19 vaccine development landscape. Nat Rev Drug Discov 2020;19:305–6.

12. Ahn DG, Shin HJ, Kim MH, et al. Current Status of Epidemiology, Diagnosis, Therapeutics, and Vaccines for Novel Coronavirus Disease 2019 (COVID-19). J Microbiol Biotechnol 2020;30:313–24.

13. Roper RL, Rehm KE. SARS vaccines: where are we? Expert Rev Vaccines 2009;8:887–98.

14. Hellerstein M. What are the roles of antibodies versus a durable, high quality T-cell response in protective immunity against SARS-CoV-2? Vaccine X 2020;6:100076.

15. Dagotto G, Yu J, Barouch DH. Approaches and Challenges in SARS-CoV-2 Vaccine Development. Cell Host Microbe 2020;28:364–70.

16. Enjuanes L, Zuñiga S, Castaño-Rodriguez C, Gutierrez-Alvarez J, Canton J, Sola I. Molecular Basis of Coronavirus Virulence and Vaccine Development. Adv Virus Res 2016;96:245–86.

17. Sariol A, Perlman S. Lessons for COVID-19 Immunity from Other Coronavirus Infections. Immunity 2020;53:248–63.

18. Du L, He Y, Zhou Y, Liu S, Zheng BJ, Jiang S. The spike protein of SARS-CoV--a target for vaccine and therapeutic development. Nat Rev Microbiol 2009;7:226–36.

19. Ou X, Liu Y, Lei X, et al. Characterization of spike glycoprotein of SARS-CoV-2 on virus entry and its immune cross-reactivity with SARS-CoV. Nat Commun 2020;11:1620.

20. Huang AT, Garcia-Carreras B, Hitchings MDT, et al. A systematic review of antibody mediated immunity to coronaviruses: kinetics, correlates of protection, and association with severity. Nat Commun 2020;11:4704.

21. Casadevall A, Joyner MJ, Pirofski LA. SARS-CoV-2 viral load and antibody responses: the case for convalescent plasma therapy. J Clin Invest 2020;130:5112–4.

22. Tang F, Quan Y, Xin ZT, et al. Lack of peripheral memory B cell responses in recovered patients with severe acute respiratory syndrome: a six-year follow-up study. J Immunol 2011;186:7264–8.

23. Kaneko N, Kuo HH, Boucau J, et al. Loss of Bcl-6-Expressing T Follicular Helper Cells and Germinal Centers in COVID-19. Cell 2020;183:143-57.e13.

24. Chen Z, John Wherry E. T cell responses in patients with COVID-19. Nat Rev Immunol 2020;20:529–36.

25. Toor SM, Saleh R, Sasidharan Nair V, Taha RZ, Elkord E. T cell responses and therapies against SARS-CoV-2 infection. Immunology 2020.

26. Ng OW, Chia A, Tan AT, et al. Memory T cell responses targeting the SARS coronavirus persist up to 11 years post-infection. Vaccine 2016;34:2008–14.

27. Channappanavar R, Fett C, Zhao J, Meyerholz DK, Perlman S. Virus-specific memory CD8 T cells provide substantial protection from lethal severe acute respiratory syndrome coronavirus infection. J Virol 2014;88:11034–44.

28. Pillet S, Couillard J, Trépanier S, et al. Immunogenicity and safety of a quadrivalent plant-derived virus like particle influenza vaccine candidate-Two randomized Phase II clinical trials in 18 to 49 and ≥50 years old adults. PLoS One 2019;14:e0216533.

29. Makarkov AI, Golizeh M, Ruiz-Lancheros E, et al. Plant-derived virus-like particle vaccines drive cross-presentation of influenza A hemagglutinin peptides by human monocyte-derived macrophages. NPJ Vaccines 2019;4:17.

30. Landry N, Pillet S, Favre D, et al. Influenza virus-like particle vaccines made in Nicotiana benthamiana elicit durable, poly-functional and cross-reactive T cell responses to influenza HA antigens. Clin Immunol 2014;154:164–77.

31. Ward BJ, Makarkov A, Séguin A, et al. Efficacy, immunogenicity, and safety of a plant-derived, quadrivalent, virus-like particle influenza vaccine in adults (18-64 years) and older adults (≥65 years): two multicentre, randomised phase 3 trials. The Lancet 2020.

32. D’Aoust MA, Couture MM, Charland N, et al. The production of hemagglutinin-based virus-like particles in plants: a rapid, efficient and safe response to pandemic influenza. Plant Biotechnol J 2010;8:607–19.

33. Grifoni A, Weiskopf D, Ramirez SI, et al. Targets of T Cell Responses to SARS-CoV-2 Coronavirus in Humans with COVID-19 Disease and Unexposed Individuals. Cell 2020;181:1489-501.e15.

34. Pillet S, Aubin É, Trépanier S, et al. Humoral and cell-mediated immune responses to H5N1 plant-made virus-like particle vaccine are differentially impacted by alum and GLA-SE adjuvants in a Phase 2 clinical trial. npj Vaccines 2018;3:3.

35. Cohet C, van der Most R, Bauchau V, et al. Safety of AS03-adjuvanted influenza vaccines: A review of the evidence. Vaccine 2019;37:3006–21.

36. Hyer R, McGuire DK, Xing B, Jackson S, Janssen R. Safety of a two-dose investigational hepatitis B vaccine, HBsAg-1018, using a toll-like receptor 9 agonist adjuvant in adults. Vaccine 2018;36:2604–11.

37. Scheiermann J, Klinman DM. Clinical evaluation of CpG oligonucleotides as adjuvants for vaccines targeting infectious diseases and cancer. Vaccine 2014;32:6377–89.

38. HEPLISAV-B®. [package insert]. Emeryville, CA: Dynavax Technologies Corporation; 2018.

39. Huang X, Yang Y. Targeting the TLR9-MyD88 pathway in the regulation of adaptive immune responses. Expert Opin Ther Targets 2010;14:787–96.

40. Givord C, Welsby I, Detienne S, et al. Activation of the endoplasmic reticulum stress sensor IRE1α by the vaccine adjuvant AS03 contributes to its immunostimulatory properties. npj Vaccines 2018;3:20.

41. van der Most RG, Clément F, Willekens J, et al. Long-Term Persistence of Cell-Mediated and Humoral Responses to A(H1N1)pdm09 Influenza Virus Vaccines and the Role of the AS03 Adjuvant System in Adults during Two Randomized Controlled Trials. Clin Vaccine Immunol 2017;24.

42. Pandemirix®. [package insert]. North Carolina. US. GlaxoSmith Kline.

43. Leroux-Roels G, Marchant A, Levy J, et al. Impact of adjuvants on CD4(+) T cell and B cell responses to a protein antigen vaccine: Results from a phase II, randomized, multicenter trial. Clin Immunol 2016;169:16–27.

44. Penn-Nicholson A, Geldenhuys H, Burny W, et al. Safety and immunogenicity of candidate vaccine M72/AS01E in adolescents in a TB endemic setting. Vaccine 2015;33:4025–34.

45. Muralidharan A, Li C, Wang L, Li X. Immunopathogenesis associated with formaldehyde-inactivated RSV vaccine in preclinical and clinical studies. Expert Rev Vaccines 2017;16:351–60.

46. Lambert PH, Ambrosino DM, Andersen SR, et al. Consensus summary report for CEPI/BC March 12-13, 2020 meeting: Assessment of risk of disease enhancement with COVID-19 vaccines. Vaccine 2020;38:4783–91.

47. Crotty S. Follicular helper CD4 T cells (TFH). Annu Rev Immunol 2011;29:621–63.

48. Zhu J. T helper 2 (Th2) cell differentiation, type 2 innate lymphoid cell (ILC2) development and regulation of interleukin-4 (IL-4) and IL-13 production. Cytokine 2015;75:14–24.

49. Ward BJ, Landry N, Trépanier S, et al. Human antibody response to N-glycans present on plant-made influenza virus-like particle (VLP) vaccines. Vaccine 2014;32:6098–106.

50. Pillet S, Aubin É, Trépanier S, et al. A plant-derived quadrivalent virus like particle influenza vaccine induces cross-reactive antibody and T cell response in healthy adults. Clin Immunol 2016;168:72–87.

## Supplementary References

1. Law B, Sturkenboom M. Safety platform for emergency vaccines (SPEAC). D2.3 Priority list of adverse events of special interest: COVID-19. Report v1.1. 2020 Mar 5.

2. Law B, Sturkenboom M. Safety platform for emergency vaccines (SPEAC). D2.3 Priority list of adverse events of special interest: COVID-19. Report v2.0. 2020a May 25.

